# Ultra-processed food intake and colorectal cancer risk in the NIH-AARP Diet and Health Study

**DOI:** 10.1101/2025.11.25.25339608

**Authors:** Leila Abar, Caitlin P. O’Connell, Hyokyoung G. Hong, Kirsten A. Herrick, Lisa Kahle, Jennifer L. Lerman, Linda M. Liao, Xuehong Zhang, Xinyuan Zhang, Longgang Zhao, Sémi Zouiouich, Rashmi Sinha, Neha Khandpur, Eurídice Martínez Steele, Erikka Loftfield

**Affiliations:** Metabolic Epidemiology Branch, Division of Cancer Epidemiology and Genetics, National Cancer Institute, National Institutes of Health, Rockville, MD; Biostatistics Branch, Division of Cancer Epidemiology and Genetics, National Cancer Institute, National Institutes of Health, Rockville, MD; Risk Factor Assessment Branch, Division of Cancer Control and Population Sciences, National Cancer Institute, National Institutes of Health, Rockville, MD; Information Management Services (IMS), Inc., Calverton, MD; Yale School of Nursing, New Haven, CT; Channing Division of Network Medicine, Brigham and Women’s Hospital and Harvard Medical School, Boston, MA; Nutrition and Metabolism Branch, International Agency for Research on Cancer, World Health Organization, Lyon, France; Division of Human Nutrition and Health, Wageningen University, Wageningen, Netherlands; Centre for Epidemiological Studies in Health and Nutrition (NUPENS), University of São Paulo, São Paulo, Brazil; Department of Nutrition, School of Public Health, University of São Paulo, São Paulo, Brazil

## Abstract

**Background:** Ultra-processed foods (UPF) account for >50% of calories consumed by US adults. Strong evidence links whole grain, fiber, calcium, and dairy intake to lower and processed meat intake to higher colorectal (CRC) risk. UPF, include some whole grain and dairy products and most processed meats. Studies of UPF intake and CRC risk are inconsistent.

**Objective:** To estimate the association between UPF intake and CRC risk as well as to evaluate the joint effect of UPF intake and diet quality with CRC risk and to estimate associations of select food groups and nutrients with CRC risk by UPF and non-UPF source .

**Methods:** US adults, aged 50-71, who participated in the NIH-AARP Diet and Health Study self-reported dietary intake using a validated food frequency questionnaire (FFQ). We assigned disaggregated FFQ items to Nova classification and categorized UPF intake (g/1000 kcal/day) into sex-specific quintiles. We used multivariable-adjusted Cox proportional hazards regression models to estimate hazard ratios (HR) and 95% confidence intervals (CI) for CRC.

**Results:** Over 20 years of follow-up, 10,075 colorectal adenocarcinoma cases were diagnosed among 461,682 participants who were cancer-free at baseline. Median UPF intake was 293 g/1000 kcal/day or 43% of daily energy intake. UPF intake was not associated with incident CRC (HR_Q5vs.Q1_=0.97; 95% CI, 0.91-1.03; *P*_trend_=.55) overall or by anatomic location (all *P*_trend_>.05). Whole grain, dairy, and calcium intake were inversely but meat intake was positively associated with CRC risk regardless of processing level.

**Conclusions:** Total UPF intake was not associated with incident CRC in this cohort of older, US adults. This may be explained, in part, by opposing effects of some UPF on CRC etiology. Our findings support current dietary guidance to consume whole grains, fiber, dairy, and calcium and avoid processed meat for CRC prevention.

## Introduction

Colorectal cancer (CRC) is the third most commonly diagnosed cancer in the US,^1^ and incidence rates among younger adults (<50 years) have increased since the mid-1990s.^2,3^ Changes in intake of established or novel dietary factors^4–6^ may contribute to increasing rates of early-onset CRC. The 2018 World Cancer Research Fund Report concluded that higher processed meat, red meat and alcohol intake are associated with higher CRC risk, while higher whole grain, fiber, dairy and calcium intake are associated with lower CRC risk.^7^ Consumption of ultra-processed foods (UPF; Nova group 4) has been associated with higher CRC risk in some but not all prospective cohort studies,^8–12^ and it remains unclear how current evidence-based dietary guidance for CRC prevention intersects with research on food processing.

Evidence-based dietary guidelines also recommend limiting intake of added sugar, sodium, and saturated fat^13–15^ and prioritizing whole foods to improve health.^16,17^ While most UPF are energy dense and nutrient poor,^13–15^ some are not (e.g., whole wheat bread). A 2023 study demonstrated that it is possible, though perhaps not practical,^18^ for dietary patterns high in UPF to receive high diet quality scores.^19^ Therefore, we sought to explore whether diet quality or food and nutrient intake impact associations of UPF intake with CRC incidence.

Our primary aim was to estimate the association between UPF intake^20^ and CRC risk. Our secondary aims were to evaluate the joint effect of UPF intake and diet quality, measured using the Healthy Eating Index-2015 (HEI-2015),^21^ with CRC risk and to estimate associations for food group (e.g., whole grain) and nutrient (e.g., fiber) intake with CRC risk by UPF and non-UPF source .

## Methods

### Study population

The design of the National Institutes of Health (NIH)-AARP Diet and Health Study has been detailed elsewhere.^22^ In brief, NIH-AARP participants, aged 50 to 71 years, were recruited in 1995-96 from among 3.5 million AARP members residing in one of six states (California, Florida, Louisiana, New Jersey, North Carolina, and Pennsylvania) or two metropolitan areas (Atlanta, GA, and Detroit, MI). Participants self-reported information on demographic, lifestyle, and other health-related characteristics via baseline questionnaire; 566,398 participants completed the questionnaire, which was considered to imply informed consent. The NIH-AARP Study was approved by the Special Studies NIH Institutional Review Board of the National Cancer Institute.

We excluded participants who had a proxy respondent (n=15,760); self-reported cancer, except for non-melanoma skin cancer (n=51,062), end-stage renal disease (n=769), or poor self-rated health (n=8,365) at baseline; had a death record for cancer without a registry-confirmed cancer (n=14,113); were caloric outliers, defined as >2 interquartile range (IQR) above the 75^th^ percentile or below the 25^th^ percentile of sex-specific Box-Cox-transformed intake (n=3,664)^23,24^; or had <1 year of follow-up (n=10,983). Our final analytic sample included 461,682 participants.

### Exposure assessment

Dietary intake was assessed using a food frequency questionnaire (FFQ) that included 124 food and beverage items, with portion sizes and 21 questions on low-fat, high-fiber foods and food preparation methods. Methods used to convert FFQ responses into USDA food codes^20^ and classify them according to Nova^25^ have been described in detail elsewhere. Briefly, FFQ line items were comprised of 3,513 individual food codes using the USDA’s 1994–1996 Continuing Survey of Food Intake of Individuals (CSFII). CSFII food codes were matched 1:1 to 8-digit USDA Food and Nutrient Database for Dietary Studies (FNDDS) food codes, representing the most commonly consumed foods and beverages at the population level, which were unfolded into component 8-digit food codes and 3,553 unique standard reference (SR) codes.^25^ Each SR code was classified according to Nova through database linkage; gram weights and energy values were each summed to the parent food code level.^25^

To estimate intake of food groups and nutrients according to Nova, we used the Food Patterns Ingredient Database (FPID), 2005-06 through 2017-18, matching each SR code to its first occurrence in FPID; 192 SR codes that could not be directly matched in FPID databases were reviewed and assigned a proxy SR code based on descriptions that included information about whether a product was ready-to-eat/drink (e.g., “14323–orange drink, canned” was assigned as “14435–orange breakfast drink, ready-to-drink”).

The estimation and validation of gram weight and energy intake according to Nova in the NIH-AARP calibration sub-study has been described previously.^25,26^ Energy-adjusted gram weight UPF intake outperformed UPF intake based on grams or energy alone. Therefore, we used the nutrient-density method to adjust for total energy,^27^ and our primary exposure variable was sex-specific quintiles of energy-adjusted UPF intake (g/1000 kcal/day). We also created sex-specific quintiles using percentage of total energy (% kcal/day) and grams (% g/day) from UPF. We decided *a priori* to adjust for alcoholic beverage intake, so Nova variables do not include grams or calories from alcoholic beverages.^28^

### Cohort follow-up and case ascertainment

Incident cancer cases were identified through cancer registry linkage in study states and three states popular for relocation (Arizona, Nevada, and Texas). Vital status was determined through National Death Index linkage. Follow-up time was defined from baseline to date of cancer diagnosis, death, relocation outside catchment area, or end of study follow-up (December 31, 2018). Exit age was calculated based on entry age and duration of follow-up.

CRC cases were identified using the International Classification of Diseases for Oncology, Third Edition (ICD-O-3). We restricted our definition to primary adenocarcinoma of the proximal colon (C180, C182, C183, C184), distal colon (C185, C186, C187), and rectum (C199, C209) using the following histology codes: 8140, 8141, 8143, 8145, 8210, 8211, 8221, 8260, 8261, 8262, 8263, 8470, 8480, 8481, and 8490. Cases with overlapping colon lesions (C188), unspecified colon cancer (C189), and large intestine not otherwise specified (C260) were censored at their date of diagnosis in subsite analyses.

### Statistical Analysis

We used Cox proportional hazards regression models, with age as the underlying time metric, to estimate hazard ratios (HR) and 95% confidence intervals (CI) for the association between sex-specific quintiles of UPF intake (g/1000 kcal/day) and CRC risk, overall and by anatomic location. We selected potential confounders based on the literature: self-reported sex, race/ethnicity, education level, smoking status (incorporating time since cessation and intensity), physical activity level, standard alcohol drink equivalents per day, family history of cancer, and self-rated health status.^9,12,29–31^ To assess a linear trend across UPF quintiles, we assigned each quintile its median value and treated it as a continuous variable. To test the proportional hazards assumption, we estimated a score test for a time-varying UPF intake.^32,33^ We observed a violation of proportional hazards assumption (P-value=.005). Therefore, we evaluated HR estimates for UPF intake and CRC risk during 5-year follow-up periods, ranging from <5 to >15 years of follow-up.

Because diet quality, nutrient intake, and body mass index (BMI) could be on the causal pathway between UPF intake and incident CRC, we decided *a priori* to run separate models. First, we further adjusted our main model for HEI-2015 (sex-specific quartiles), calcium intake (mg/1000 kcal/day), and fiber (g/1000 kcal/day) intake. Then, we further adjusted our main model for BMI status.

In secondary analyses, we tested for effect modification, on the multiplicative scale, by diet quality, sex, and BMI status using the likelihood ratio test to compare models with and without an interaction term for UPF and the variable of interest. We present HR estimates for the joint-associations of UPF intake (sex-specific quintiles) and diet quality (>80 “good”, 51-80 “needs improvement”, and “<51 “poor”), using a common reference group, defined as the lowest quintile of UPF intake and a “good” HEI-2015 score, which we hypothesized to be the lowest risk group. We present HR estimates stratified by sex and BMI category.

We calculated mutually adjusted HR estimates for continuous intake of whole grains (servings/1000 kcal/day), dietary fiber (scaled to 10 g/1000 kcal/day), dairy (servings/1000 kcal/day), dietary calcium (scaled to 300 mg/1000 kcal/day), or meat (servings/1000 kcal/day) from UPF and non-UPF sources. To compare our analysis directly with current literature, we also ran a compositional substitution model, including ^34^ to assess the effect of substituting 10% of grams from minimally processed food (MPF; Nova group1) for 10% of grams from UPF; this model included the relative intakes (percentage of grams) from Nova group 1, 2, and 3, and left out the relative intake from Nova group 4, such that the relative risk estimate for Nova group 1 can be interpreted as substituting Nova group 1 for Nova group 4 while keeping Nova groups 2 and 3 constant.^35^ Finally, we evaluated associations for sex-specific quintiles, based on percentage energy (% kcal/day) or grams from UPF (% g/day), with incident CRC, overall and by anatomic location.

All analyses were performed using R version 4.4.1 (R Core Team, 2024). R Foundation for Statistical Computing, Vienna, Austria. A two-sided P-value <.05 was considered statistically significant.

## Results

Our analytic sample included 274,269 men and 187,413 women (N=461,682). Median (IQR) UPF intake was 293 (198-466) g/1000 kcal/day, corresponding to 42.6% (35.9-49.6) of total daily energy intake. With median follow-up of 18.8 years (IQR 9.7-22.5), 10,075 CRC cases were ascertained; 4,787 were proximal colon, 2,633 were distal colon, and 2,389 were rectal cancer.

Participants in the highest quintile (Q5) of UPF intake were younger (60.7 vs. 63.8 years; **Table 1 & Supplementary Table 1**) than those in the lowest quintile (Q1). They were less likely to be Asian (0.5% vs. 3.0%), never smokers (32.9% vs. 36.0%), or college graduates (34.4% vs. 41.8%), but more likely to be alcohol non-drinkers (30.7% vs. 21.5%) or have obesity (29.6% vs. 15.2%). They were also less likely to exercise ≥3 times/week (40.6% vs. 49.7%) or rate their health as excellent (14.2% vs. 21.4%).

**Table 1.** Baseline characteristics of study participants, by sex-specific quintiles of ultra-processed food intake (g/1000 kcal/day), in the NIH-AARP Diet and Health Study (N=461,682)

| Characteristic, N (%) | UPF Q1<br>(n=92337) | UPF Q2<br>(n=92336) | UPF Q3<br>(n=92336) | UPF Q4<br>(n=92336) | UPF Q5<br>(n=92337) |
| --- | --- | --- | --- | --- | --- |
| **Range UPF, g/1000 kcal/day, male** | 3-187 | >187-254 | >254-350 | >350-525 | >525-10043 |
| **Range UPF, g/1000 kcal/day, female** | 10-177 | >177-241 | >241-343 | >343-530 | >530-12740 |
| **Median UPF, g/1000 kcal/day (IQR), male** | 155 (134-172) | 218 (202-235) | 298 (275-323) | 419 (382-465) | 727 (606-959) |
| **Median UPF, g/1000 kcal/day (IQR), female** | 147 (127-163) | 207 (191-223) | 285 (262-313) | 418 (378-467) | 757 (619-1026) |
| **Median age, years (IQR)** | 63.8 (59.0-67.3) | 63.4 (58.5-67.0) | 62.7 (57.9-66.6) | 61.9 (57.3-66.1) | 60.7 (56.1-65.3) |
| **Sex, n (%)** |  |  |  |  |  |
| Male | 54854 (59.4%) | 54854 (59.4%) | 54853 (59.4%) | 54854 (59.4%) | 54854 (59.4%) |
| Female | 37483 (40.6%) | 37482 (40.6%) | 37483 (40.6%) | 37482 (40.6%) | 37483 (40.6%) |
| **Race/ethnicity, n (%)** |  |  |  |  |  |
| American Indian/Alaskan Native | 254 (0.3%) | 218 (0.2%) | 246 (0.3%) | 242 (0.3%) | 298 (0.3%) |
| Asian | 2740 (3.0%) | 1155 (1.3%) | 917 (1.0%) | 628 (0.7%) | 416 (0.5%) |
| Hispanic | 2463 (2.7%) | 1988 (2.2%) | 1743 (1.9%) | 1423 (1.5%) | 1221 (1.3%) |
| Non-Hispanic Black | 3275 (3.5%) | 3025 (3.3%) | 3637 (3.9%) | 3754 (4.1%) | 3922 (4.2%) |
| Non-Hispanic White | 81869 (88.7%) | 84657 (91.7%) | 84634 (91.7%) | 85149 (92.2%) | 85221 (92.3%) |
| Pacific Islander | 200 (0.2%) | 104 (0.1%) | 104 (0.1%) | 80 (0.1%) | 64 (0.1%) |
| Unknown | 1536 (1.7%) | 1189 (1.3%) | 1055 (1.1%) | 1060 (1.1%) | 1195 (1.3%) |
| **Never smoker, n (%)** | 33247 (36.0%) | 33732 (36.5%) | 33820 (36.6%) | 33245 (36.0%) | 30420 (32.9%) |
| **College graduate, n (%)** | 38610 (41.8%) | 38487 (41.7%) | 36425 (39.4%) | 34301 (37.1%) | 31702 (34.3%) |
| **Exercises $\geq 3$ times a week, n (%)** | 45863 (49.7%) | 44994 (48.7%) | 43444 (47.1%) | 41456 (44.9%) | 38447 (41.6%) |
| **Alcohol non-drinkers, n (%)** | 19819 (21.5%) | 19019 (20.6%) | 20243 (21.9%) | 22589 (24.5%) | 28362 (30.7%) |
| **BMI $\geq 30$ kg/m<sup>2</sup>, n (%)** | 14045 (15.2%) | 15948 (17.3%) | 18784 (20.3%) | 21773 (23.6%) | 27328 (29.6%) |
| **Excellent self-rated health, n (%)** | 19804 (21.4%) | 17524 (19.0%) | 15975 (17.3%) | 14617 (15.8%) | 13112 (14.2%) |
| **Family history of cancer, n (%)** | 44444 (48.1%) | 45407 (49.2%) | 45069 (48.8%) | 44931 (48.7%) | 44322 (48.0%) |
| **Median total energy intake, kcal/day (IQR)** | 1547 (1165-2025) | 1622 (1241-2106) | 1671 (1268-2172) | 1615 (1246-2126) | 1520 (1126-2020) |
| **Median total HEI-2015 score (IQR)** | 70.9 (64.1-76.7) | 69.9 (63.0-75.7) | 68.9 (62.1-74.7) | 67.6 (60.6-73.6) | 65.6 (58.3-72.0) |
| **Median dietary calcium, mg/1000 kcal/day (IQR)** | 417 (327-555) | 419 (335-542) | 416 (332-535) | 403 (321-519) | 405 (315-528) |
| **Median dietary fiber, g/1000 kcal/day (IQR)** | 11.8 (9.3-14.9) | 11.4 (9.2-14.0) | 10.8 (8.8-13.4) | 10.4 (8.4-12.9) | 10.0 (7.8-12.6) |

Abbreviations: UPF, Ultra-processed food; HEI-2015, Healthy Eating Index-2015; IQR, interquartile range; BMI, body mass index

In multivariable-adjusted models, no associations were observed between UPF (g/1000 kcal/day) quintiles and CRC risk overall (HR_Q5vs.Q1_=0.97; 95% CI=0.91 to 1.03; *P*_trend_=.55) or by subsite (proximal colon: HR_Q5vs.Q1_=1.02, 0.93 to 1.12, *P*_trend_=.20; distal colon: HR_Q5vs.Q1_=0.94, 0.83 to 1.06, *P*_trend_=.64; rectum: HR_Q5vs.Q1_=0.91, 0.80 to 1.03, *P*_trend_=.93; **Table 2**). Adjusting for HEI-2015 score and nutrient intake (HR_Q5vs.Q1_=0.94; 0.88 to 1.00; *P*_trend_=.69) or BMI status (HR_Q5vs.Q1_=0.94; 0.89 to 1.01; *P*_trend_=.86) did not appreciably alter HR estimates (<10% change).

**Table 2.**
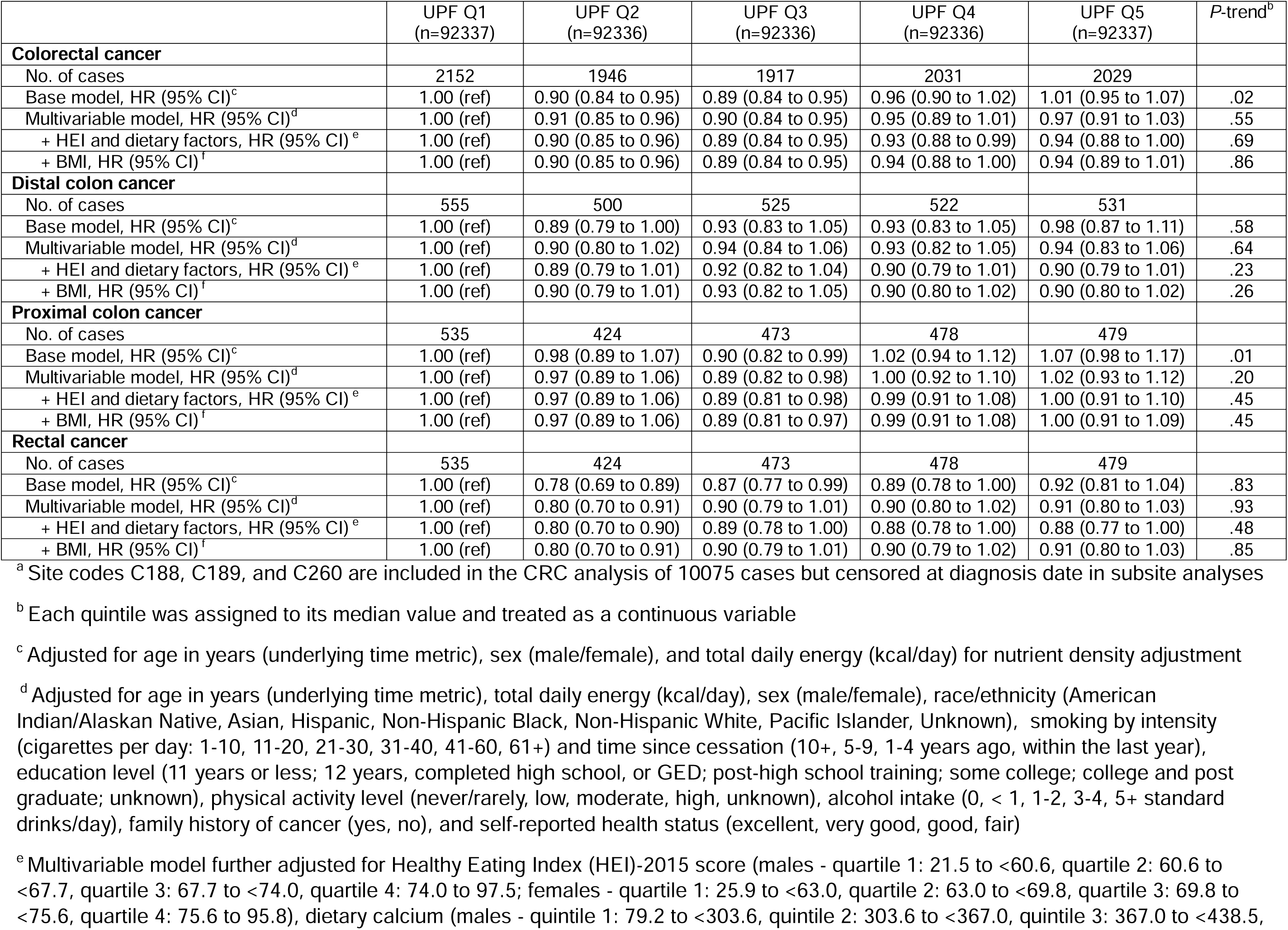
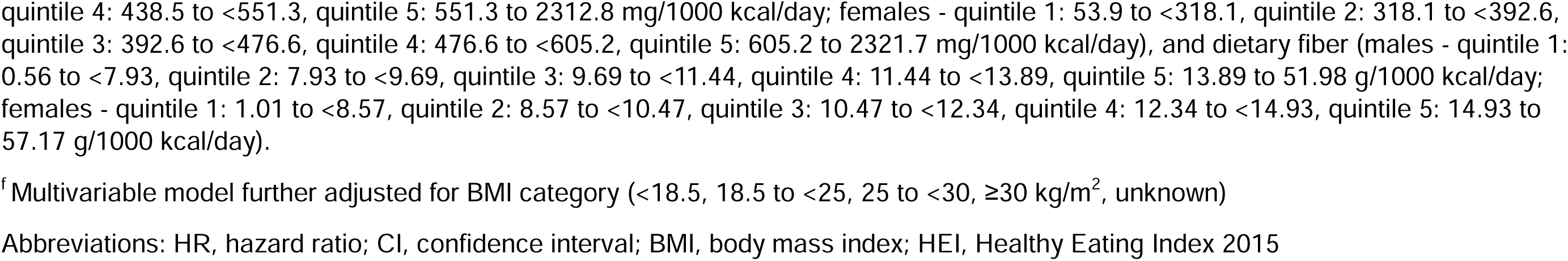
Association of ultra-processed food intake (sex-specific quintiles of nutrient-adjusted g/1000 kcal/day) with colorectal cancer risk overall and by anatomic locationa in the NIH-AARP Diet and Health Study (N=461,682)

|  | UPF Q1<br>(n=92337) | UPF Q2<br>(n=92336) | UPF Q3<br>(n=92336) | UPF Q4<br>(n=92336) | UPF Q5<br>(n=92337) | P-trend<sup>b</sup> |
| --- | --- | --- | --- | --- | --- | --- |
| **Colorectal cancer** |  |  |  |  |  |  |
| No. of cases | 2152 | 1946 | 1917 | 2031 | 2029 |  |
| Base model, HR (95% CI)<sup>c</sup> | 1.00 (ref) | 0.90 (0.84 to 0.95) | 0.89 (0.84 to 0.95) | 0.96 (0.90 to 1.02) | 1.01 (0.95 to 1.07) | .02 |
| Multivariable model, HR (95% CI)<sup>d</sup> | 1.00 (ref) | 0.91 (0.85 to 0.96) | 0.90 (0.84 to 0.95) | 0.95 (0.89 to 1.01) | 0.97 (0.91 to 1.03) | .55 |
| + HEI and dietary factors, HR (95% CI)<sup>e</sup> | 1.00 (ref) | 0.90 (0.85 to 0.96) | 0.89 (0.84 to 0.95) | 0.93 (0.88 to 0.99) | 0.94 (0.88 to 1.00) | .69 |
| + BMI, HR (95% CI)<sup>f</sup> | 1.00 (ref) | 0.90 (0.85 to 0.96) | 0.89 (0.84 to 0.95) | 0.94 (0.88 to 1.00) | 0.94 (0.89 to 1.01) | .86 |
| **Distal colon cancer** |  |  |  |  |  |  |
| No. of cases | 555 | 500 | 525 | 522 | 531 |  |
| Base model, HR (95% CI)<sup>c</sup> | 1.00 (ref) | 0.89 (0.79 to 1.00) | 0.93 (0.83 to 1.05) | 0.93 (0.83 to 1.05) | 0.98 (0.87 to 1.11) | .58 |
| Multivariable model, HR (95% CI)<sup>d</sup> | 1.00 (ref) | 0.90 (0.80 to 1.02) | 0.94 (0.84 to 1.06) | 0.93 (0.82 to 1.05) | 0.94 (0.83 to 1.06) | .64 |
| + HEI and dietary factors, HR (95% CI)<sup>e</sup> | 1.00 (ref) | 0.89 (0.79 to 1.01) | 0.92 (0.82 to 1.04) | 0.90 (0.79 to 1.01) | 0.90 (0.79 to 1.01) | .23 |
| + BMI, HR (95% CI)<sup>f</sup> | 1.00 (ref) | 0.90 (0.79 to 1.01) | 0.93 (0.82 to 1.05) | 0.90 (0.80 to 1.02) | 0.90 (0.80 to 1.02) | .26 |
| **Proximal colon cancer** |  |  |  |  |  |  |
| No. of cases | 535 | 424 | 473 | 478 | 479 |  |
| Base model, HR (95% CI)<sup>c</sup> | 1.00 (ref) | 0.98 (0.89 to 1.07) | 0.90 (0.82 to 0.99) | 1.02 (0.94 to 1.12) | 1.07 (0.98 to 1.17) | .01 |
| Multivariable model, HR (95% CI)<sup>d</sup> | 1.00 (ref) | 0.97 (0.89 to 1.06) | 0.89 (0.82 to 0.98) | 1.00 (0.92 to 1.10) | 1.02 (0.93 to 1.12) | .20 |
| + HEI and dietary factors, HR (95% CI)<sup>e</sup> | 1.00 (ref) | 0.97 (0.89 to 1.06) | 0.89 (0.81 to 0.98) | 0.99 (0.91 to 1.08) | 1.00 (0.91 to 1.10) | .45 |
| + BMI, HR (95% CI)<sup>f</sup> | 1.00 (ref) | 0.97 (0.89 to 1.06) | 0.89 (0.81 to 0.97) | 0.99 (0.91 to 1.08) | 1.00 (0.91 to 1.09) | .45 |
| **Rectal cancer** |  |  |  |  |  |  |
| No. of cases | 535 | 424 | 473 | 478 | 479 |  |
| Base model, HR (95% CI)<sup>c</sup> | 1.00 (ref) | 0.78 (0.69 to 0.89) | 0.87 (0.77 to 0.99) | 0.89 (0.78 to 1.00) | 0.92 (0.81 to 1.04) | .83 |
| Multivariable model, HR (95% CI)<sup>d</sup> | 1.00 (ref) | 0.80 (0.70 to 0.91) | 0.90 (0.79 to 1.01) | 0.90 (0.80 to 1.02) | 0.91 (0.80 to 1.03) | .93 |
| + HEI and dietary factors, HR (95% CI)<sup>e</sup> | 1.00 (ref) | 0.80 (0.70 to 0.90) | 0.89 (0.78 to 1.00) | 0.88 (0.78 to 1.00) | 0.88 (0.77 to 1.00) | .48 |
| + BMI, HR (95% CI)<sup>f</sup> | 1.00 (ref) | 0.80 (0.70 to 0.91) | 0.90 (0.79 to 1.01) | 0.90 (0.79 to 1.02) | 0.91 (0.80 to 1.03) | .85 |

<sup>a</sup> Site codes C188, C189, and C260 are included in the CRC analysis of 10075 cases but censored at diagnosis date in subsite analyses
<sup>b</sup> Each quintile was assigned to its median value and treated as a continuous variable
<sup>c</sup> Adjusted for age in years (underlying time metric), sex (male/female), and total daily energy (kcal/day) for nutrient density adjustment
<sup>d</sup> Adjusted for age in years (underlying time metric), total daily energy (kcal/day), sex (male/female), race/ethnicity (American Indian/Alaskan Native, Asian, Hispanic, Non-Hispanic Black, Non-Hispanic White, Pacific Islander, Unknown), smoking by intensity (cigarettes per day: 1-10, 11-20, 21-30, 31-40, 41-60, 61+) and time since cessation (10+, 5-9, 1-4 years ago, within the last year), education level (11 years or less; 12 years, completed high school, or GED; post-high school training; some college; college and post graduate; unknown), physical activity level (never/rarely, low, moderate, high, unknown), alcohol intake (0, < 1, 1-2, 3-4, 5+ standard drinks/day), family history of cancer (yes, no), and self-reported health status (excellent, very good, good, fair)
<sup>e</sup> Multivariable model further adjusted for Healthy Eating Index (HEI)-2015 score (males - quartile 1: 21.5 to <60.6, quartile 2: 60.6 to <67.7, quartile 3: 67.7 to <74.0, quartile 4: 74.0 to 97.5; females - quartile 1: 25.9 to <63.0, quartile 2: 63.0 to <69.8, quartile 3: 69.8 to <75.6, quartile 4: 75.6 to 95.8), dietary calcium (males - quintile 1: 79.2 to <303.6, quintile 2: 303.6 to <367.0, quintile 3: 367.0 to <438.5,
quintile 4: 438.5 to <551.3, quintile 5: 551.3 to 2312.8 mg/1000 kcal/day; females - quintile 1: 53.9 to <318.1, quintile 2: 318.1 to <392.6, quintile 3: 392.6 to <476.6, quintile 4: 476.6 to <605.2, quintile 5: 605.2 to 2321.7 mg/1000 kcal/day), and dietary fiber (males - quintile 1: 0.56 to <7.93, quintile 2: 7.93 to <9.69, quintile 3: 9.69 to <11.44, quintile 4: 11.44 to <13.89, quintile 5: 13.89 to 51.98 g/1000 kcal/day; females - quintile 1: 1.01 to <8.57, quintile 2: 8.57 to <10.47, quintile 3: 10.47 to <12.34, quintile 4: 12.34 to <14.93, quintile 5: 14.93 to 57.17 g/1000 kcal/day).
<sup>f</sup> Multivariable model further adjusted for BMI category (<18.5, 18.5 to <25, 25 to <30, ≥30 kg/m<sup>2</sup>, unknown)
Abbreviations: HR, hazard ratio; CI, confidence interval; BMI, body mass index; HEI, Healthy Eating Index 2015

In estimating the joint-effect of UPF intake and diet quality, no statistical evidence of a multiplicative interaction (*P*_heterogeneity_=.11) was observed. HR estimates across joint UPF-HEI categories were inconsistent; compared to being in the lowest quintile of UPF intake with good diet quality, being in the 2^nd^ (HR=1.28; 1.01 to 1.50) or 5^th^ (HR=1.22; 1.01 to 1.48) quintile with poor diet quality was associated with higher CRC risk (**Figure 1**). No evidence of effect modification by sex (*P*_heterogeneity_=.30; **Supplementary tables 2 & 3**) or BMI status (*P*_heterogeneity_=.38; **Supplementary table 4**) was observed. HR estimates during 5-year follow-up periods were generally similar (**Table 3**).

**Figure 1.**
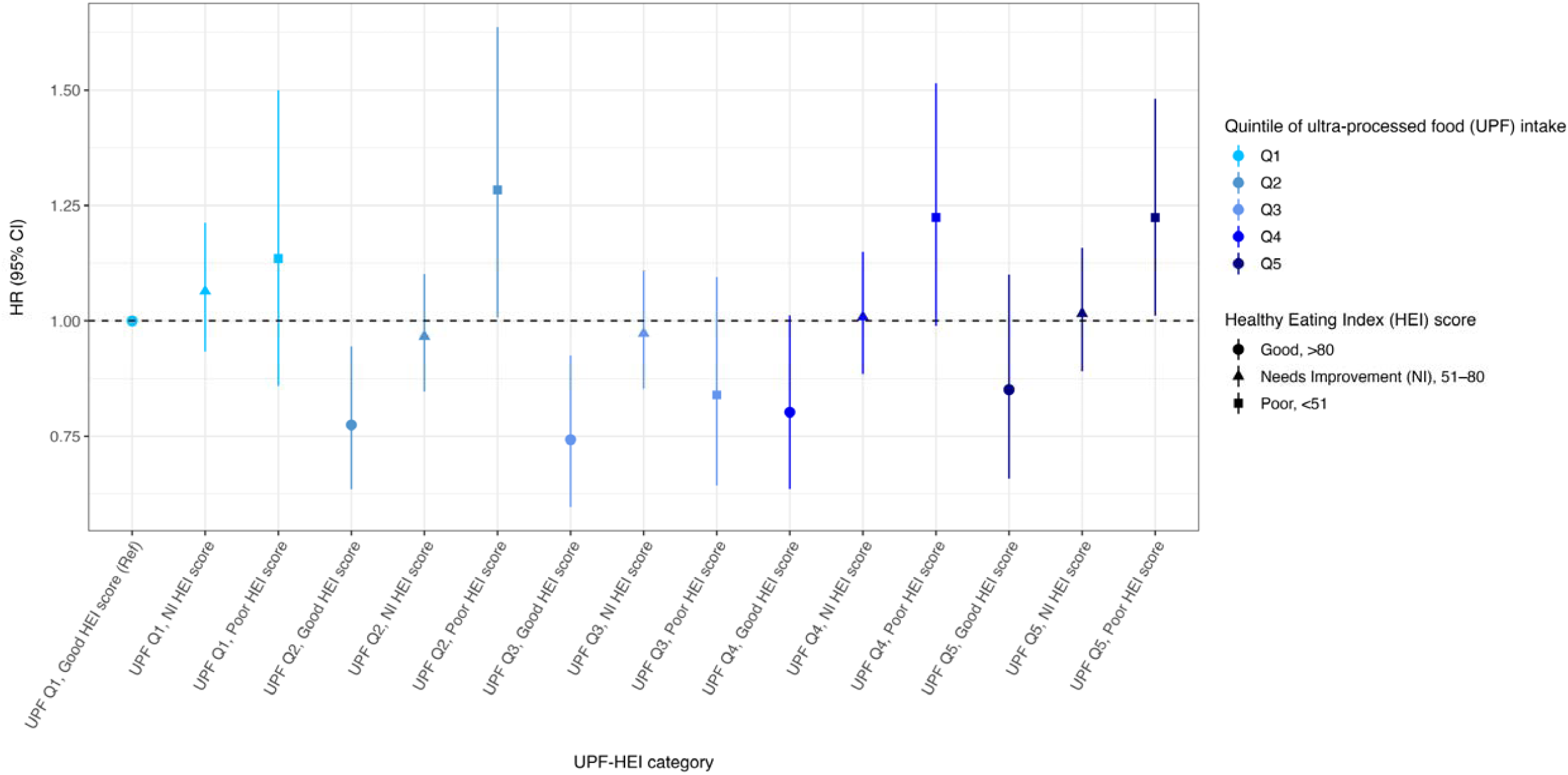
Associations for joint effect of ultra-processed food (UPF) intake (g/1000 kcal/day), defined using sex-specific quintiles, and diet quality, measured using the Healthy Eating Index (HEI)-2015, with colorectal cancer risk in the NIH-AARP Diet and Health Study cohort (N=461,682) ^a^ Hazard ratios (HR) and 95% confidence intervals (CI) are estimated using a Cox proportional hazard regression model adjusted for age in years (underlying time metric), total daily energy (kcal/day), sex (male/female), race/ethnicity (American Indian/Alaskan Native, Asian, Hispanic, Non-Hispanic Black, Non-Hispanic White, Pacific Islander, Unknown), smoking by intensity (cigarettes per day: 1-10, 11-20, 21-30, 31-40, 41-60, 61+) and time since cessation (10+, 5-9, 1-4 years ago, within the last year), education level (11 years or less; 12 years, completed high school, or GED; post-high school training; some college; college and post graduate; unknown), physical activity level (never/rarely, low, moderate, high, unknown), alcohol intake (0, < 1, 1-2, 3-4, 5+ standard drinks/day), family history of cancer (yes, no), and self-reported health status (excellent, very good, good, fair). UPF Q1 and HEI-2015 score>80 serves as the common reference group. Likelihood ratio test comparing models with and without UPF*HEI-2015 interaction term: P=.11.

**Table 3.**
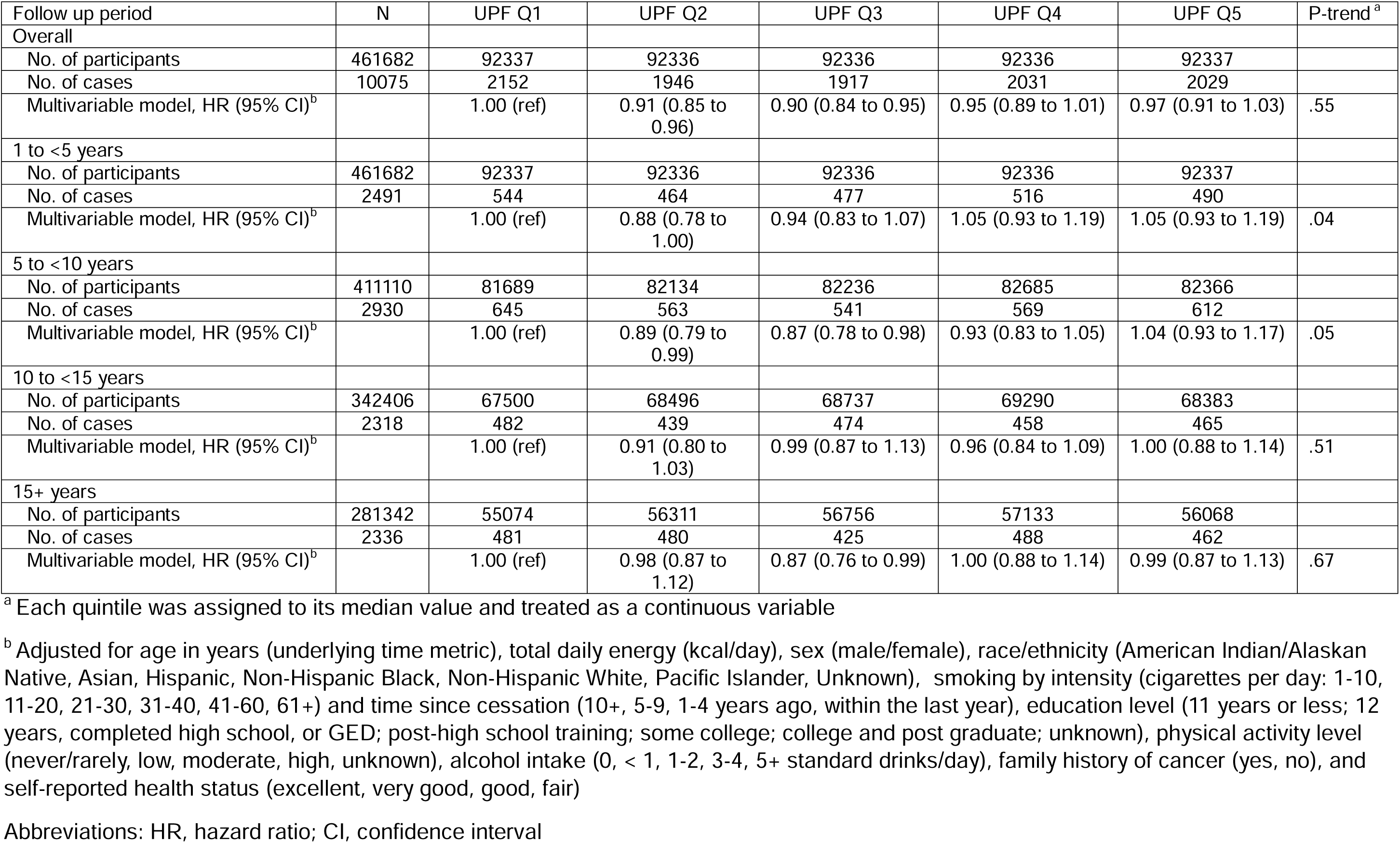
Association of ultra-processed food intake (sex-specific quintiles of nutrient-adjusted g/1000 kcal/day) with colorectal cancer risk according to follow-up time (1to <5 y, 5 to <10 y, 10 to <15 y, ≥15 y) in the NIH-AARP Diet and Health Study (N=461,682)

In our analysis, 82.8%, 33.5%, 31.7% and 23.5% of whole grain, dietary fiber, dietary calcium, and dairy intake, respectively, as well as 74.3% of processed meat intake came from UPF sources (**Supplementary table 5**). In mutually-adjusted models, dietary calcium (HR_UPF_=0.77, 0.69 to 0.86; HR_non-UPF_=0.90, 0.87 to 0.93) and dairy (HR_UPF_=0.75, 0.62 to 0.91; HR_non-UPF_=0.90, 0.87 to 0.94) intake from UPF and non-UPF were independently associated with lower CRC risk, and UPF and non-UPF meat intake (HR_UPF_=1.08, 1.02 to 1.14; HR_non-UPF_=1.04, 1.01 to 1.06) were independently associated with higher CRC risk. Whole grain (HR=0.87, 0.82 to 0.92) and fiber (HR=0.72, 0.64 to 0.80) intake from UPF were inversely associated with CRC risk but their non-UPF counterparts in the model were not (**Figure 2**).

**Figure 2.**
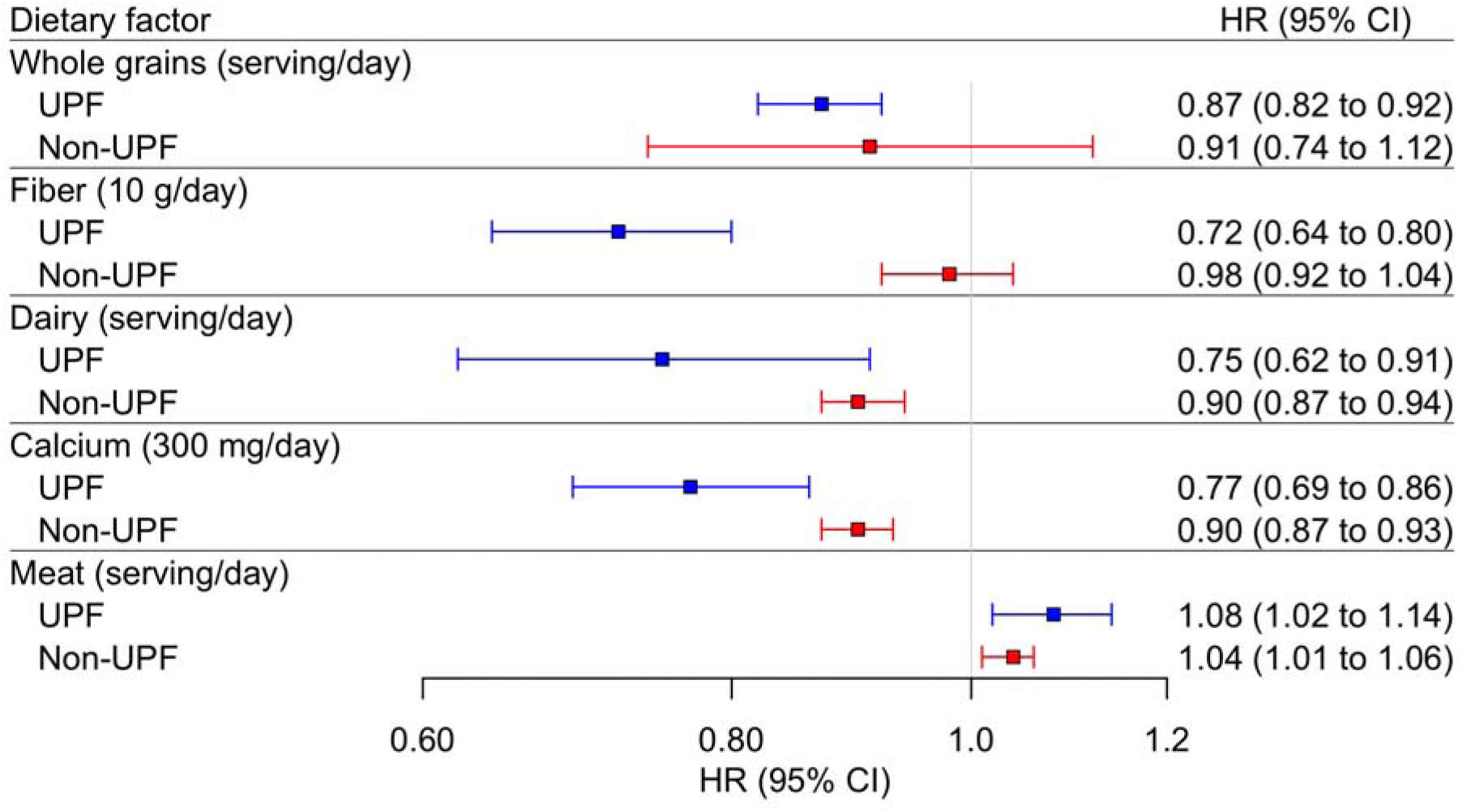
Mutually adjusted associations for select dietary factors with colorectal cancer risk by Nova classification of food source (UPF or non-UPF) in the NIH-AARP Diet and Health Study (N=461,682) Hazard ratios (HR) and 95% confidence intervals (CI) are estimated using a Cox proportional hazard regression model adjusted for age in years (underlying time metric), total daily energy (kcal/day), sex (male/female), race/ethnicity (American Indian/Alaskan Native, Asian, Hispanic, Non-Hispanic Black, Non-Hispanic White, Pacific Islander, Unknown), smoking by intensity (cigarettes per day: 1-10, 11-20, 21-30, 31-40, 41-60, 61+) and time since cessation (10+, 5-9, 1-4 years ago, within the last year), education level (11 years or less; 12 years, completed high school, or GED; post-high school training; some college; college and post graduate; unknown), physical activity level (never/rarely, low, moderate, high, unknown), alcohol intake (0, < 1, 1-2, 3-4, 5+ standard drinks/day), family history of cancer (yes, no), and self-reported health status (excellent, very good, good, fair). The model accounts for mutual adjustments between sources from both non-UPF (Nova 1-3) and UPF (Nova 4). Abbreviations: UPF, ultra-processed food; HR, hazard ratio; CI, confidence interval

Following multivariable adjustment (main model), substituting 10% of grams from UPF with 10% from MPF was associated with a 2% lower risk of CRC (HR=0.98, 0.96 to 0.99). UPF intake, defined using percentage energy or grams (**Supplementary tables 6 & 7**), generally yielded similar null HR estimates. In contrast, those in the highest, compared to the lowest, quintile of percentage energy from UPF had lower rectal cancer risk (HR_Q5vs.Q1_=0.86, 0.76 to 0.98; *P*_trend_=0.05); adjusting for diet quality, fiber, and calcium strengthened the inverse association (HR_Q5vs.Q1_=0.76, 0.67 to 0.88; *P*_trend_<.001).

## Discussion

In the NIH-AARP Diet and Health Study, with an analytic cohort of 461,682 US adults who were followed for cancer for more than 20 years, we found little evidence of an association between total UPF intake and CRC risk. Interestingly, we found that in NIH-AARP, UPF contributed to intake of dietary factors that have been associated with lower CRC risk,^7,36–38^ namely whole grains (82.8% from UPF), dietary fiber (33.5%), dairy (23.5%), and dietary calcium (31.7%), as well as to intake of processed meat (74.3% from UPF), an established risk factor for CRC.^39^ Finally, we found that CRC associations for intake of whole grains, dietary fiber, dairy, dietary calcium, and processed meat reflected current dietary guidance for CRC prevention, regardless of food processing level.^7^

An umbrella review,^40^ which synthesized results from epidemiologic studies, ^8,9,12,41–43^ concluded that the quality of evidence for an association between UPF and CRC was very low.^40^ Prospective studies on UPF intake and CRC risk have been mixed. In the French NutriNet-Santé cohort, UPF intake (% grams/day) was associated with higher CRC risk; however, the association, which was based on 153 CRC cases, was not statistically significant.^9^ Similarly, analyses of multiple cancer types in the UK Biobank (UKB) and European Prospective Investigation into Cancer (EPIC) found no association between UPF intake (% grams/day) and CRC risk.^12,35^ An analysis of data from three US cohorts of health professionals found that higher UPF intake (servings/day) was associated with higher CRC risk in men, but not in women,^8^ whereas, a subsequent analysis of data from the EPIC study showed a 6% higher CRC risk for each 10% increase in UPF intake (% g/day) in women but not in men.^11^ In EPIC, Nahas et al. also found that substituting 10% of grams from UPF with 10% from MPF was associated with a 6% lower risk of CRC (HR=0.94, 0.90 to 0.97).^11^ We conducted a similar analysis and found that in NIH-AARP the same substitution was association with a 2% lower risk of CRC (HR=0.98, 0.96 to 0.99).

One plausible explanation for varying results is that the composition of diets high in UPF intake differs across populations. For example, higher intake of ultra-processed meat products (e.g., mean 36.2 g/d in EPIC vs. 20.1 g/d in NIH-AARP; **Supplementary table 8**) and lower intake of some ultra-processed breakfast cereals (e.g., mean 5.0 g/d in EPIC vs. 15.2 g/d in NIH-AARP) and breads (e.g., mean 43.6 g/d in EPIC vs. 57.3 g/d in NIH-AARP),^35^ which serve a sources of whole grain intake, could contribute to observed differences across studies or across strata (e.g., sex) within a study and explain why the HR estimate for replacing 10% grams from UPF with MPF was similar in direction to the EPIC analysis but weaker in magnitude in NIH-AARP.

Poor diet quality has been associated with higher CRC risk in the NIH-AARP Study.^44–47^ Herein, we found that poor diet quality was associated with elevated CRC risk in both lower and higher quintiles of UPF intake. Looking specifically at nutrient and food group intake, we found that dairy and calcium intake were associated with lower CRC risk, but meat intake was associated with higher CRC risk in models containing intake estimates (e.g., calcium) from both from UPF and non-UPF. In contrast, we observed inverse associations for whole grain and fiber intake from UPF, but not non-UPF, sources. This could be explained by the fact that <20% of estimated whole grain intake came from non-UPF sources, indicating that these foods were consumed by fewer people or in small amounts and likely resulting in less precise HR estimates. For fiber, although most intake came from non-UPF sources, the association with lower CRC risk in NIH-AARP is for fiber from whole grains,^36^ which come mostly from UPF sources in our study, likely explaining the non-significant inverse association for non-UPF fiber.

Our study has limitations. First, the FFQ and databases that our study relied on were designed to measure nutrient intake not to assess food processing level. Individuals who reported eating yogurt, for example, could have consumed flavored or plain yogurt, which fall under different Nova groups, but this level of granularity was not captured directly. The approach used to estimate nutrient values of FFQ items in NIH-AARP uses weighting to account for varying levels of nutrients across commonly consumed food products. Since details on processing level were not specifically captured in CSFII, a similar weighting approach for applying Nova classification relies on assumptions about processing levels, which may limit the exposure differences that can be captured between individuals and attenuate associations toward the null. Still, we previously demonstrated that the NIH-AARP FFQ performs reasonably well, as compared with two 24h dietary recalls, for estimating intake according to the Nova system^26^ with attenuation factors comparable to those for nutrients and food groups.^22^ Exposure misclassification may have been compounded in food group and nutrient analyses since intake estimation relied on an additional level of classification; thus, these analyses should be viewed as exploratory and interpreted with caution. Still, we expect measurement error to be non-differential with respect to CRC, likely attenuating associations toward the null, owing to the prospective study design. Participants were older, mostly white, US adults who were recruited in the mid-1990s, and UPF intake was measured only once at baseline. Consequently, our findings may not generalize to younger more diverse populations or reflect the everchanging US food supply. Given substantial changes in UPF availability and consumption over the past few decades,^48^ our estimates likely underestimate current UPF intake levels, which may also contribute to observed null findings. Limited variation across quintiles of UPF intake may also contribute to our null findings. UPF often contain food additives that may negatively impact gut health.^49–51^ For example, emulsifiers have been found to disrupt the intestinal mucosal barrier, leading to dysbiosis, increased gut permeability, and low-grade inflammation—factors linked to metabolic syndrome and CRC.^52–54^ However, we were unable to estimate associations with food additives due to the absence of ingredient level data.

Strengths of our study include its large size and extended follow-up for cancer outcomes, which afforded sufficient case numbers to evaluate associations by tumor location and diet quality. Furthermore, our method for disaggregating FFQ items to assign Nova classification^26^ not only enhances reliability of our UPF measure but also allowed us to estimate intake of nutrients and food groups by Nova classification of the food source.

In conclusion, we found that UPF intake was not associated with CRC risk in a large cohort of older US adults. Understanding how current evidence-based dietary guidance for CRC prevention intersects with research on food processing is critical. Our results support a role for whole grain, fiber, dairy and calcium intake in CRC prevention that was not diminished by food processing level. However, poor diet quality is a risk factor for CRC,^44–47^ and dietary patterns that limit intake of added sugars, sodium, saturated fat, and refined grains and prioritize whole foods lower risk of chronic disease-related mortality.^55^ In addition, substituting 10% of grams from UPF with 10% from MPF was associated with a 2% lower risk of CRC. Future studies are needed to replicate and extend our findings, and novel methods for measuring UPF and additive intake are needed to elucidate what aspects of UPF are associated with CRC risk.

## Acknowledgements

Cancer incidence data from the Atlanta metropolitan area were collected by the Georgia Center for Cancer Statistics, Department of Epidemiology, Rollins School of Public Health, Emory University, Atlanta, Georgia. Cancer incidence data from California were collected by the California Cancer Registry, California Department of Public Health’s Cancer Surveillance and Research Branch, Sacramento, California. Cancer incidence data from the Detroit metropolitan area were collected by the Michigan Cancer Surveillance Program, Community Health Administration, Lansing, Michigan. The Florida cancer incidence data used in this report were collected by the Florida Cancer Data System (Miami, Florida) under contract with the Florida Department of Health, Tallahassee, Florida. The views expressed herein are solely those of the authors and do not necessarily reflect those of the FCDC or FDOH. Cancer incidence data from Louisiana were collected by the Louisiana Tumor Registry, Louisiana State University Health Sciences Center School of Public Health, New Orleans, Louisiana. Cancer incidence data from New Jersey were collected by the New Jersey State Cancer Registry, The Rutgers Cancer Institute of New Jersey, New Brunswick, New Jersey. Cancer incidence data from North Carolina were collected by the North Carolina Central Cancer Registry, Raleigh, North Carolina. Cancer incidence data from Pennsylvania were supplied by the Division of Health Statistics and Research, Pennsylvania Department of Health, Harrisburg, Pennsylvania. The Pennsylvania Department of Health specifically disclaims responsibility for any analyses, interpretations or conclusions. Cancer incidence data from Arizona were collected by the Arizona Cancer Registry, Division of Public Health Services, Arizona Department of Health Services, Phoenix, Arizona. Cancer incidence data from Texas were collected by the Texas Cancer Registry, Cancer Epidemiology and Surveillance Branch, Texas Department of State Health Services, Austin, Texas. Cancer incidence data from Nevada were collected by the Nevada Central Cancer Registry, Division of Public and Behavioral Health, State of Nevada Department of Health and Human Services, Carson City, Nevada.

We are indebted to the participants in the NIH-AARP Diet and Health Study for their outstanding cooperation. We also thank Sigurd Hermansen and Kerry Grace Morrissey from Westat for study outcomes ascertainment and management and Leslie Carroll at Information Management Services for data support and analysis.

## Data availability

Data are maintained by the National Cancer Institute, Division of Cancer Epidemiology and Genetics and are available upon approval of a proposal submitted to the NIH-AARP Diet and Health Study Steering Committee. For more information visit https://www.nihaarpstars.com/.

