## Supplemental material for "Ultra-processed food intake and colorectal cancer risk in the NIH-AARP Diet and Health Study"

Supplementary Table 1. Baseline characteristics of the study participants, by nutrient-adjusted ultra-processed food (UPF) intake^a^ (N=461,682)

| **Characteristic, N (%)**^b^ | **Quintile 1 (N=92337)** | **Quintile 2 (N=92336)** | **Quintile 3 (N=92336)** | **Quintile 4 (N=92336)** | **Quintile 5 (N=92337)** |
| --- | --- | --- | --- | --- | --- |
| **Median grams of UPF**, g/day (IQR) | 226 (162-308) | 346 (261-456) | 491 (364-649) | 672 (524-905) | 1160 (889-1787) |
| **Median grams of UPF**, g/1000 kcal/day (IQR) | 152 (131-168) | 213 (198-230) | 293 (270-319) | 419 (380-466) | 738 (611-986) |
| **Median age**, years (IQR) | 63.8 (59.0-67.3) | 63.4 (58.5-67.0) | 62.7 (57.9-66.6) | 61.9 (57.3-66.1) | 60.7 (56.1-65.3) |
| **Sex** |  |  |  |  |  |
| Male | 54854 (59.4%) | 54854 (59.4%) | 54853 (59.4%) | 54854 (59.4%) | 54854 (59.4%) |
| Female | 37483 (40.6%) | 37482 (40.6%) | 37483 (40.6%) | 37482 (40.6%) | 37483 (40.6%) |
| **Race/ethnicity** |  |  |  |  |  |
| American Indian/Alaskan Native | 254 (0.3%) | 218 (0.2%) | 246 (0.3%) | 242 (0.3%) | 298 (0.3%) |
| Asian | 2740 (3.0%) | 1155 (1.3%) | 917 (1.0%) | 628 (0.7%) | 416 (0.5%) |
| Hispanic | 2463 (2.7%) | 1988 (2.2%) | 1743 (1.9%) | 1423 (1.5%) | 1221 (1.3%) |
| Non-Hispanic Black | 3275 (3.5%) | 3025 (3.3%) | 3637 (3.9%) | 3754 (4.1%) | 3922 (4.2%) |
| Non-Hispanic White | 81869 (88.7%) | 84657 (91.7%) | 84634 (91.7%) | 85149 (92.2%) | 85221 (92.3%) |
| Pacific Islander | 200 (0.2%) | 104 (0.1%) | 104 (0.1%) | 80 (0.1%) | 64 (0.1%) |
| Unknown | 1536 (1.7%) | 1189 (1.3%) | 1055 (1.1%) | 1060 (1.1%) | 1195 (1.3%) |
| **Smoking status** |  |  |  |  |  |
| Never smoker | 33247 (36.0%) | 33732 (36.5%) | 33820 (36.6%) | 33245 (36.0%) | 30420 (32.9%) |
| Former smoker | 43359 (47.0%) | 45451 (49.2%) | 45613 (49.4%) | 45608 (49.4%) | 46066 (49.9%) |
| Current smoker | 11870 (12.9%) | 9767 (10.6%) | 9554 (10.3%) | 10207 (11.1%) | 12313 (13.3%) |
| Unknown | 3861 (4.2%) | 3386 (3.7%) | 3349 (3.6%) | 3276 (3.5%) | 3538 (3.8%) |
| **Education** |  |  |  |  |  |
| less than 12 years | 5209 (5.6%) | 4897 (5.3%) | 5177 (5.6%) | 5345 (5.8%) | 5947 (6.4%) |
| 12 years, completed high school, GED | 15892 (17.2%) | 16980 (18.4%) | 17940 (19.4%) | 19372 (21.0%) | 19939 (21.6%) |
| post-high school training | 8399 (9.1%) | 8510 (9.2%) | 9112 (9.9%) | 9569 (10.4%) | 9716 (10.5%) |
| some college | 21184 (22.9%) | 20953 (22.7%) | 21162 (22.9%) | 21320 (23.1%) | 22288 (24.1%) |
| college and postgraduate | 38610 (41.8%) | 38487 (41.7%) | 36425 (39.4%) | 34301 (37.1%) | 31702 (34.3%) |
| Unknown | 3043 (3.3%) | 2509 (2.7%) | 2520 (2.7%) | 2429 (2.6%) | 2745 (3.0%) |
| **Physical activity** |  |  |  |  |  |
| Never/Rarely | 15062 (16.3%) | 14340 (15.5%) | 15014 (16.3%) | 16012 (17.3%) | 19164 (20.8%) |
| Low (1-3 times a month) | 11382 (12.3%) | 12015 (13.0%) | 12396 (13.4%) | 13078 (14.2%) | 13959 (15.1%) |
| Moderate (1-2 times per week) | 18784 (20.3%) | 20065 (21.7%) | 20635 (22.3%) | 20881 (22.6%) | 19782 (21.4%) |
| High (≥3 times a week) | 45863 (49.7%) | 44994 (48.7%) | 43444 (47.1%) | 41456 (44.9%) | 38447 (41.6%) |
| Unknown | 1246 (1.3%) | 922 (1.0%) | 847 (0.9%) | 909 (1.0%) | 985 (1.1%) |
| **Alcohol intake category** |  |  |  |  |  |
| 0 drinks per day | 19819 (21.5%) | 19019 (20.6%) | 20243 (21.9%) | 22589 (24.5%) | 28362 (30.7%) |
| <1 drink per day | 44867 (48.6%) | 49473 (53.6%) | 51281 (55.5%) | 51496 (55.8%) | 47955 (51.9%) |
| 1-2 drinks per day | 17457 (18.9%) | 16498 (17.9%) | 14531 (15.7%) | 12623 (13.7%) | 10011 (10.8%) |
| 3-4 drinks per day | 4795 (5.2%) | 3797 (4.1%) | 3202 (3.5%) | 2984 (3.2%) | 3087 (3.3%) |
| ≥5 drinks per day | 5399 (5.8%) | 3549 (3.8%) | 3079 (3.3%) | 2644 (2.9%) | 2922 (3.2%) |
| **Body mass index category** |  |  |  |  |  |
| <18.5 kg/m^2^ | 1272 (1.4%) | 961 (1.0%) | 865 (0.9%) | 771 (0.8%) | 735 (0.8%) |
| 18.5 to <25 kg/m^2^ | 38386 (41.6%) | 35442 (38.4%) | 31508 (34.1%) | 27596 (29.9%) | 22976 (24.9%) |
| 25 to <30 kg/m^2^ | 36059 (39.1%) | 37886 (41.0%) | 39147 (42.4%) | 40184 (43.5%) | 39218 (42.5%) |
| ≥ 30 kg/m^2^ | 14045 (15.2%) | 15948 (17.3%) | 18784 (20.3%) | 21773 (23.6%) | 27328 (29.6%) |
| Unknown | 2575 (2.8%) | 2099 (2.3%) | 2032 (2.2%) | 2012 (2.2%) | 2080 (2.3%) |
| **Family history of cancer** |  |  |  |  |  |
| No | 42935 (46.5%) | 42398 (45.9%) | 42592 (46.1%) | 42723 (46.3%) | 43137 (46.7%) |
| Yes | 44444 (48.1%) | 45407 (49.2%) | 45069 (48.8%) | 44931 (48.7%) | 44322 (48.0%) |
| Unknown | 4958 (5.4%) | 4531 (4.9%) | 4675 (5.1%) | 4682 (5.1%) | 4878 (5.3%) |
| **Self-reported health status** |  |  |  |  |  |
| Excellent | 19804 (21.4%) | 17524 (19.0%) | 15975 (17.3%) | 14617 (15.8%) | 13112 (14.2%) |
| Very good | 33852 (36.7%) | 34650 (37.5%) | 33758 (36.6%) | 33078 (35.8%) | 30318 (32.8%) |
| Good | 28927 (31.3%) | 30284 (32.8%) | 32167 (34.8%) | 33576 (36.4%) | 34864 (37.8%) |
| Fair | 8271 (9.0%) | 8522 (9.2%) | 9084 (9.8%) | 9776 (10.6%) | 12619 (13.7%) |
| Unknown | 1483 (1.6%) | 1356 (1.5%) | 1352 (1.5%) | 1289 (1.4%) | 1424 (1.5%) |
| **Median total HEI-2015 score**, IQR | 70.9 (64.1-76.7) | 69.9 (63.0-75.7) | 68.9 (62.1-74.7) | 67.6 (60.6-73.6) | 65.6 (58.3-72.0) |
| **Median total energy intake (kcal)**, IQR | 1550 (1165-2025) | 1620 (1241-2106) | 1670 (1268-2172) | 1620 (1246-2126) | 1520 (1126-2020) |
| **Median dietary calcium mg/1000 kcal/day**, IQR | 417 (327-555) | 419 (335-542) | 416 (332-535) | 403 (321-519) | 405 (315-528) |
| **Median dietary fiber g/1000 kcal/day**, IQR | 11.8 (9.3-14.9) | 11.4 (9.2-14.0) | 10.8 (8.8-13.4) | 10.4 (8.4-12.9) | 10.0 (7.8-12.6) |

Abbreviations: UPF, Ultra-processed food; HEI-2015, Healthy Eating Index-2015; IQR, interquartile range; BMI, body mass index

^a^Quintiles of grams classified as Nova group 4 nutrient adjusted.

^b^Column percentages might not add up to exactly 100% due to rounding.

Supplementary Table 2. Association of ultra-processed food intake (quintiles of nutrient-adjusted g/1000 kcal/day) with colorectal cancer risk in male participants in the NIH-AARP Diet and Health Study (N=274,269)

| Cancer type | Overall no. of cases^a^ | Q1 HR (95% CI)  (N=54854) | Q2 HR (95% CI)  (N=54854) | Q3 HR (95% CI)  (N=54853) | Q4 HR (95% CI)  (N=54854) | Q5 HR (95% CI)  (N=54854) | *P* for trend^b^ |
| --- | --- | --- | --- | --- | --- | --- | --- |
| Colorectal cancer |  |  |  |  |  |  |  |
| No. of cases | 6414 | 1380 | 1216 | 1244 | 1267 | 1307 |  |
| Base model hazard ratio (95% CI)^c^ |  | 1.00 (ref) | 0.87 (0.81 to 0.94) | 0.90 (0.84 to 0.97) | 0.93 (0.86 to 1.00) | 1.00 (0.92 to 1.07) | .11 |
| Multivariable-adjusted hazard ratio^d^ (95% CI) |  | 1.00 (ref) | 0.88 (0.82 to 0.95) | 0.91 (0.84 to 0.98) | 0.93 (0.86 to 1.00) | 0.97 (0.90 to 1.04) | .56 |
| Adjusted for HEI and dietary factors^e^ |  | 1.00 (ref) | 0.88 (0.81 to 0.95) | 0.90 (0.83 to 0.97) | 0.90 (0.84 to 0.98) | 0.94 (0.86 to 1.01) | .77 |
| Adjusted for BMI^f^ |  | 1.00 (ref) | 0.88 (0.82 to 0.95) | 0.90 (0.84 to 0.98) | 0.92 (0.85 to 0.99) | 0.95 (0.88 to 1.02) | .99 |
| Distal colon |  |  |  |  |  |  |  |
| No. of cases | 1774 | 370 | 338 | 375 | 337 | 354 |  |
| Base model hazard ratio (95% CI)^c^ |  | 1.00 (ref) | 0.90 (0.78 to 1.04) | 1.00 (0.87 to 1.16) | 0.90 (0.78 to 1.04) | 0.97 (0.84 to 1.13) | .98 |
| Multivariable-adjusted hazard ratio^d^ (95% CI) |  | 1.00 (ref) | 0.92 (0.79 to 1.06) | 1.01 (0.87 to 1.17) | 0.90 (0.77 to 1.04) | 0.93 (0.80 to 1.08) | .43 |
| Adjusted for HEI and dietary factors^e^ |  | 1.00 (ref) | 0.90 (0.78 to 1.05) | 0.98 (0.85 to 1.14) | 0.86 (0.74 to 1.00) | 0.88 (0.76 to 1.03) | .13 |
| Adjusted for BMI^f^ |  | 1.00 (ref) | 0.91 (0.79 to 1.06) | 0.99 (0.86 to 1.15) | 0.88 (0.75 to 1.02) | 0.90 (0.77 to 1.04) | .17 |
| Proximal colon |  |  |  |  |  |  |  |
| No. of cases | 2807 | 585 | 557 | 516 | 567 | 582 |  |
| Base model hazard ratio (95% CI)^c^ |  | 1.00 (ref) | 0.95 (0.85 to 1.07) | 0.90 (0.80 to 1.01) | 1.00 (0.89 to 1.12) | 1.09 (0.97 to 1.22) | .019 |
| Multivariable-adjusted hazard ratio^d^ (95% CI) |  | 1.00 (ref) | 0.95 (0.85 to 1.07) | 0.90 (0.79 to 1.01) | 0.99 (0.88 to 1.11) | 1.05 (0.93 to 1.18) | .10 |
| Adjusted for HEI and dietary factors^e^ |  | 1.00 (ref) | 0.95 (0.84 to 1.07) | 0.89 (0.79 to 1.00) | 0.98 (0.87 to 1.10) | 1.03 (0.92 to 1.16) | .18 |
| Adjusted for BMI^f^ |  | 1.00 (ref) | 0.95 (0.84 to 1.07) | 0.89 (0.79 to 1.00) | 0.98 (0.87 to 1.10) | 1.02 (0.91 to 1.15) | .26 |
| Rectal |  |  |  |  |  |  |  |
| No. of cases | 1660 | 375 | 291 | 329 | 330 | 335 |  |
| Base model hazard ratio (95% CI)^c^ |  | 1.00 (ref) | 0.76 (0.66 to 0.89) | 0.87 (0.75 to 1.00) | 0.87 (0.75 to 1.01) | 0.91 (0.78 to 1.05) | .91 |
| Multivariable-adjusted hazard ratio^d^ (95% CI) |  | 1.00 (ref) | 0.79 (0.67 to 0.92) | 0.89 (0.77 to 1.03) | 0.89 (0.77 to 1.03) | 0.91 (0.78 to 1.05) | .96 |
| Adjusted for HEI and dietary factors^e^ |  | 1.00 (ref) | 0.78 (0.66 to 0.90) | 0.87 (0.75 to 1.01) | 0.86 (0.74 to 1.00) | 0.86 (0.74 to 1.00) | .45 |
| Adjusted for BMI^f^ |  | 1.00 (ref) | 0.79 (0.67 to 0.92) | 0.89 (0.77 to 1.04) | 0.89 (0.77 to 1.04) | 0.91 (0.78 to 1.06) | .98 |

Abbreviations: HR denotes hazard ratio; CI denotes confidence interval; BMI is body mass index, calculated as weight in kilograms divided by height in meters squared; HEI is Healthy Eating Index 2015.

^a^Cases with cancer sites of C188, C189, and C260 are included in the overall analysis of 10075 cases but are censored at diagnosis date in the anatomic location analysis.

^b^Each quintile was assigned to its median value and treated as a continuous variable.

^c^Adjusted for age in years (underlying time metric), sex (male/female), and total daily energy (kcal/day) for nutrient density adjustment.

^d^Multivariable estimates were adjusted for age in years (underlying time metric), total daily energy (kcal/day), race/ethnicity (American Indian/Alaskan Native, Asian, Hispanic, Non-Hispanic Black, Non-Hispanic White, Pacific Islander, Unknown), smoking by intensity (cigarettes per day: 1-10, 11-20, 21-30, 31-40, 41-60, >60) and time since cessation (≥ 10 years ago, 5-9 years ago, 1-4 years ago, within the last year), education level (11 years or less; 12 years, completed high school, or GED; post-high school training; some college; college and post graduate; unknown), physical activity level (never/rarely, low, moderate, high, unknown), alcohol intake (0 drinks/day, < 1 drink/day, 1-2 drinks/day, 3-4 drinks/day, ≥ 5 drinks/day), family history of cancer (yes/no), and self-reported health status (excellent, very good, good, fair).

^e^Multivariable model further adjusted for Healthy Eating Index (HEI)-2015 score quartiles (quartile 1: 21.5 to <60.6, quartile 2: 60.6 to <67.7, quartile 3: 67.7 to <74.0, quartile 4: 74.0 to 97.5), nutrient-density adjusted dietary calcium quintiles (quintile 1: 79.2 to <303.6, quintile 2: 303.6 to <367.0, quintile 3: 367.0 to <438.5, quintile 4: 438.5 to <551.3, quintile 5: 551.3 to 2312.8 mg/1000 kcal/day), and nutrient-density adjusted dietary fiber quintiles (quintile 1: 0.56 to <7.93, quintile 2: 7.93 to <9.69, quintile 3: 9.69 to <11.44, quintile 4: 11.44 to <13.89, quintile 5: 13.89 to 51.98 g/1000 kcal/day).

^f^Multivariable model further adjusted for body mass index category (<18.5 kg/m2, 18.5 to <25 kg/m2, 25 to <30 kg/m2, ≥30 kg/m2, unknown).

Supplementary Table 3. Association of ultra-processed food intake (quintiles of nutrient-adjusted g/1000 kcal/day) with colorectal cancer risk in female participants in the NIH-AARP Diet and Health Study (N=187,413)

| Cancer type | Overall no. of cases^a^ | Q1 HR (95% CI)  (N=37483) | Q2 HR (95% CI)  (N=37482) | Q3 HR (95% CI)  (N=37483) | Q4 HR (95% CI)  (N=37482) | Q5 HR (95% CI)  (N=37483) | *P* for trend^b^ |
| --- | --- | --- | --- | --- | --- | --- | --- |
| Colorectal cancer |  |  |  |  |  |  |  |
| No. of cases | 3661 | 772 | 730 | 673 | 764 | 722 |  |
| Base model hazard ratio (95% CI)^c^ |  | 1.00 (ref) | 0.94 (0.85 to 1.04) | 0.87 (0.79 to 0.97) | 1.02 (0.92 to 1.13) | 1.03 (0.93 to 1.14) | .10 |
| Multivariable-adjusted hazard ratio^d^ (95% CI) |  | 1.00 (ref) | 0.95 (0.86 to 1.06) | 0.88 (0.79 to 0.98) | 1.01 (0.91 to 1.11) | 0.97 (0.88 to 1.08) | .78 |
| Adjusted for HEI and dietary factors^e^ |  | 1.00 (ref) | 0.96 (0.86 to 1.06) | 0.88 (0.79 to 0.98) | 1.00 (0.90 to 1.11) | 0.95 (0.86 to 1.06) | .88 |
| Adjusted for BMI^f^ |  | 1.00 (ref) | 0.95 (0.86 to 1.05) | 0.87 (0.79 to 0.97) | 0.99 (0.90 to 1.10) | 0.95 (0.86 to 1.06) | .92 |
| Distal colon |  |  |  |  |  |  |  |
| No. of cases | 859 | 185 | 162 | 150 | 185 | 177 |  |
| Base model hazard ratio (95% CI)^c^ |  | 1.00 (ref) | 0.87 (0.70 to 1.07) | 0.80 (0.64 to 0.99) | 1.00 (0.82 to 1.23) | 1.00 (0.81 to 1.23) | .32 |
| Multivariable-adjusted hazard ratio^d^ (95% CI) |  | 1.00 (ref) | 0.89 (0.72 to 1.10) | 0.82 (0.66 to 1.02) | 1.01 (0.82 to 1.24) | 0.96 (0.78 to 1.18) | .72 |
| Adjusted for HEI and dietary factors^e^ |  | 1.00 (ref) | 0.90 (0.72 to 1.11) | 0.82 (0.66 to 1.02) | 0.99 (0.80 to 1.22) | 0.94 (0.76 to 1.16) | .89 |
| Adjusted for BMI^f^ |  | 1.00 (ref) | 0.89 (0.72 to 1.09) | 0.81 (0.65 to 1.01) | 0.98 (0.80 to 1.21) | 0.92 (0.75 to 1.14) | .97 |
| Proximal colon |  |  |  |  |  |  |  |
| No. of cases | 1980 | 409 | 415 | 365 | 412 | 379 |  |
| Base model hazard ratio (95% CI)^c^ |  | 1.00 (ref) | 1.01 (0.88 to 1.16) | 0.90 (0.78 to 1.04) | 1.05 (0.92 to 1.21) | 1.05 (0.92 to 1.21) | .26 |
| Multivariable-adjusted hazard ratio^d^ (95% CI) |  | 1.00 (ref) | 1.02 (0.89 to 1.16) | 0.90 (0.78 to 1.03) | 1.03 (0.90 to 1.19) | 0.99 (0.86 to 1.14) | .93 |
| Adjusted for HEI and dietary factors^e^ |  | 1.00 (ref) | 1.02 (0.89 to 1.17) | 0.90 (0.78 to 1.04) | 1.02 (0.89 to 1.18) | 0.97 (0.84 to 1.12) | .76 |
| Adjusted for BMI^f^ |  | 1.00 (ref) | 1.01 (0.88 to 1.16) | 0.89 (0.78 to 1.03) | 1.03 (0.89 to 1.18) | 0.98 (0.85 to 1.13) | .98 |
| Rectal |  |  |  |  |  |  |  |
| No. of cases | 729 | 160 | 133 | 144 | 148 | 144 |  |
| Base model hazard ratio (95% CI)^c^ |  | 1.00 (ref) | 0.83 (0.66 to 1.04) | 0.89 (0.71 to 1.12) | 0.93 (0.74 to 1.16) | 0.94 (0.75 to 1.18) | .85 |
| Multivariable-adjusted hazard ratio^d^ (95% CI) |  | 1.00 (ref) | 0.85 (0.67 to 1.07) | 0.92 (0.73 to 1.15) | 0.94 (0.75 to 1.18) | 0.92 (0.73 to 1.16) | .90 |
| Adjusted for HEI and dietary factors^e^ |  | 1.00 (ref) | 0.85 (0.68 to 1.08) | 0.93 (0.74 to 1.17) | 0.95 (0.75 to 1.19) | 0.93 (0.73 to 1.17) | .91 |
| Adjusted for BMI^f^ |  | 1.00 (ref) | 0.85 (0.67 to 1.07) | 0.91 (0.73 to 1.15) | 0.92 (0.74 to 1.16) | 0.90 (0.71 to 1.13) | .71 |

Abbreviations: HR denotes hazard ratio; CI denotes confidence interval; BMI is body mass index, calculated as weight in kilograms divided by height in meters squared; HEI is Healthy Eating Index 2015.

^a^Cases with cancer sites of C188, C189, and C260 are included in the overall analysis of 3661 cases but are censored at diagnosis date in the anatomic location analysis.

^b^Each quintile was assigned to its median value and treated as a continuous variable.

^c^Adjusted for age in years (underlying time metric) and total daily energy (kcal/day) for nutrient density adjustment.

^d^Multivariable estimates were adjusted for age in years (underlying time metric), total daily energy (kcal/day), race/ethnicity (American Indian/Alaskan Native, Asian, Hispanic, Non-Hispanic Black, Non-Hispanic White, Pacific Islander, Unknown), smoking by intensity (cigarettes per day: 1-10, 11-20, 21-30, 31-40, 41-60, 61+) and time since cessation (10+ years ago, 5-9 years ago, 1-4 years ago, within the last year), education level (11 years or less; 12 years, completed high school, or GED; post-high school training; some college; college and post graduate; unknown), physical activity level (never/rarely, low, moderate, high, unknown), alcohol intake (0 drinks/day, < 1 drink/day, 1-2 drinks/day, 3-4 drinks/day, 5 or more drinks/day), family history of cancer (yes/no), self-reported health status (excellent, very good, good, fair), and menopausal hormone therapy use (never, former, current, unknown).

^e^Multivariable model further adjusted for Healthy Eating Index (HEI)-2015 score quartiles (quartile 1: 25.9 to <63.0, quartile 2: 63.0 to <69.8, quartile 3: 69.8 to <75.6, quartile 4: 75.6 to 95.8), nutrient-density adjusted dietary calcium quintiles (quintile 1: 53.9 to <318.1, quintile 2: 318.1 to <392.6, quintile 3: 392.6 to <476.6, quintile 4: 476.6 to <605.2, quintile 5: 605.2 to 2321.7 mg/1000 kcal/day), and nutrient-density adjusted dietary fiber quintiles (quintile 1: 1.01 to <8.57, quintile 2: 8.57 to <10.47, quintile 3: 10.47 to <12.34, quintile 4: 12.34 to <14.93, quintile 5: 14.93 to 57.17 g/1000 kcal/day).

^f^Multivariable model further adjusted for body mass index category (<18.5 kg/m2, 18.5 to <25 kg/m2, 25 to <30 kg/m2, ≥30 kg/m2, unknown).

Supplementary table 4. Association of ultra-processed food intake (quintiles of nutrient-adjusted g/1000 kcal/day) with colorectal cancer risk stratified by body mass index category

| BMI category | Q1 HR^a^ (95% CI) | Q2 HR^a^ (95% CI) | Q3 HR^a^ (95% CI) | Q4 HR^a^ (95% CI) | Q5 HR^a^ (95% CI) |
| --- | --- | --- | --- | --- | --- |
| < 18.5 kg/m2 | 1.00 | 0.78 (0.42 to 1.44) | 1.22 (0.69 to 2.13) | 0.69 (0.35 to 1.39) | 0.91 (0.47 to 1.77) |
| 18.5 to <25 kg/m2 | 1.00 | 0.89 (0.81 to 0.99) | 0.85 (0.76 to 0.94) | 0.90 (0.81 to 1.01) | 0.91 (0.81 to 1.02) |
| 25 to <30 kg/m2 | 1.00 | 0.93 (0.85 to 1.02) | 0.90 (0.81 to 0.99) | 0.97 (0.89 to 1.07) | 0.92 (0.83 to 1.01) |
| ≥ 30 kg/m2 | 1.00 | 0.87 (0.76 to 1.01) | 0.91 (0.79 to 1.04) | 0.89 (0.78 to 1.02) | 0.98 (0.86 to 1.11) |

Abbreviations: HR denotes hazard ratio; CI denotes confidence interval; BMI is body mass index, calculated as weight in kilograms divided by height in meters squared

^a^Multivariable estimates were adjusted for age in years (underlying time metric), total daily energy (kcal/day), sex (male/female), race/ethnicity (American Indian/Alaskan Native, Asian, Hispanic, Non-Hispanic Black, Non-Hispanic White, Pacific Islander, Unknown), smoking by intensity (cigarettes per day: 1-10, 11-20, 21-30, 31-40, 41-60, 61+) and time since cessation (10+ years ago, 5-9 years ago, 1-4 years ago, within the last year), education level (11 years or less; 12 years, completed high school, or GED; post-high school training; some college; college and post graduate; unknown), physical activity level (never/rarely, low, moderate, high, unknown), alcohol intake (0 drinks/day, < 1 drink/day, 1-2 drinks/day, 3-4 drinks/day, 5 or more drinks/day), family history of cancer (yes/no), and self-reported health status (excellent, very good, good, fair, unknown).

Supplementary table 5. Absolute intakes and relative contributions of subgroups to ultra-processed intake (Nova group 4) in NIH-AARP (N=461,682)

| Subgroup | Mean (SD) g/day | Mean (SD) % within Nova group 4 (% g/day) | Mean (SD) % within total diet (% g/day) | Mean (SD) kcal/day | Mean (SD) % within Nova group 4 (% kcal/day) | Mean (SD) % within total diet (% kcal/day) |
| --- | --- | --- | --- | --- | --- | --- |
| Reconstituted meat or fish products | 20.1 (21.9) | 4.0 (4.3) | 0.8 (0.8) | 48.9 (50.8) | 6.5 (5.3) | 2.8 (2.4) |
| Bread | 57.3 (43.2) | 12.3 (10.1) | 2.2 (1.8) | 161.4 (120.0) | 22.0 (12.0) | 9.4 (5.8) |
| Baked goods and candy^a^ | 34.5 (35.4) | 7.0 (6.8) | 1.3 (1.3) | 118.8 (134.1) | 14.8 (10.8) | 6.6 (5.7) |
| Frozen desserts and milk drinks^b^ | 46.9 (58.1) | 9.4 (11.2) | 1.8 (2.2) | 60.6 (74.2) | 8.2 (8.8) | 3.5 (3.9) |
| Breakfast cereals | 15.2 (16.1) | 3.6 (4.7) | 0.6 (0.7) | 50.4 (56.1) | 7.5 (8.4) | 3.1 (3.5) |
| Salty snacks and French fries^c^ | 15.4 (17.2) | 3.0 (3.4) | 0.6 (0.7) | 63.2 (73.7) | 8.2 (7.2) | 3.5 (3.4) |
| Ready to eat meals^d^ | 6.7 (9.0) | 1.5 (2.1) | 0.3 (0.3) | 8.6 (11.7) | 1.3 (1.7) | 0.5 (0.6) |
| Instant and canned soups | 19.6 (23.7) | 4.4 (5.4) | 0.7 (0.8) | 12.3 (14.9) | 1.8 (2.2) | 0.7 (0.8) |
| Sauces, dressings, and gravies | 32.3 (27.3) | 7.1 (6.6) | 1.2 (1.0) | 79.3 (68.8) | 11.3 (8.3) | 4.7 (3.5) |
| Carbonated soft drinks | 259.3 (469.4) | 27.5 (25.9) | 8.6 (12.3) | 36.2 (109.0) | 3.8 (8.7) | 1.9 (4.9) |
| Other sweetened beverages | 140.2 (289.3) | 16.8 (21.1) | 4.7 (7.9) | 24.5 (74.1) | 2.9 (6.2) | 1.3 (3.2) |
| Other group 4 | 16.6 (12.5) | 3.6 (3.1) | 0.6 (0.5) | 84.6 (67.0) | 11.6 (7.2) | 4.9 (3.1) |

^a^ Baked goods and candy combine Nova subgroups 24 (cakes, cookies, and pies), 26 (desserts), and 29 (sweet snacks) from Martinez Steele et al.^1^

^b^ Frozen desserts and milk drinks combine Nova subgroups 25 (ice cream and ice pops) and 37 (milk drinks).

^c^ Salty snacks and French fries combine Nova subgroups 28 (salty snacks) and 33 (French fries and other potato products).

^d^ Ready to eat meals combine Nova subgroups 30 (frozen and shelf-stable meals), 31 (pizza) and 32 (sandwiches and hamburgers on buns).

Supplementary table 6. Dietary calcium, dairy, whole grains, fiber, and meat (%) by Nova group in the NIH-AARP Diet and Health Study (N=461,682)

| Nutrient/Food group (%), mean (SD) | Nova 1 | Nova 2 | Nova 3 | Nova 4 |
| --- | --- | --- | --- | --- |
| Dietary Calcium | 55.5 (15.5) | 1.6 (2.9) | 11.3 (8.0) | 31.7 (12.4) |
| Dairy | 60.1 (25.7) | 0.9 (4.9) | 15.5 (14.5) | 23.5 (17.3) |
| Whole Grain | 17.1 (15.8) | 0 (0) | 0 (0) | 82.8 (16.0) |
| Fiber | 58.3 (12.7) | 0.2 (0.2) | 8.0 (4.6) | 33.5 (12.9) |
| Red meat | 98.4 (4.6) | 0 (0) | 0.02 (0.1) | 1.4 (1.7) |
| Processed meat | 0 (0) | 0 (0) | 25.5 (17.8) | 74.3 (18.1) |
| Total meat | 71.6 (14.0) | 0 (0) | 37.6 (39.9) | 17.4 (12.4) |

Nova 1=Unprocessed or minimally processed foods; Nova 2=Processed culinary ingredients; Nova 3=Processed foods; Nova 4=Ultra-processed foods. Abbreviation: SD = standard deviation.

Supplementary Table 7. Association of UPF intake (sex-specific quintiles of % kcal/day) with colorectal cancer risk in the NIH-AARP Diet and Health Study (N=461,682)

| Cancer type | N^a^ | Q1 HR (95% CI)  (N=92337) | Q2 HR (95% CI)  (N=92336) | Q3 HR (95% CI)  (N=92336) | Q4 HR (95% CI)  (N=92336) | Q5 HR (95% CI)  (N=92337) | *P* for trend^b^ |
| --- | --- | --- | --- | --- | --- | --- | --- |
| Colorectal cancer |  |  |  |  |  |  |  |
| No. of cases | 10075 | 2002 | 1967 | 2006 | 2003 | 2097 |  |
| Base model hazard ratio (95% CI)^c^ |  | 1.00 (ref) | 0.99 (0.93 to 1.05) | 1.00 (0.94 to 1.06) | 1.00 (0.94 to 1.06) | 1.06 (1.00 to 1.13) | .05 |
| Multivariable-adjusted hazard ratio^d^ (95% CI) |  | 1.00 (ref) | 0.97 (0.91 to 1.04) | 0.98 (0.92 to 1.04) | 0.97 (0.91 to 1.03) | 0.99 (0.93 to 1.06) | .84 |
| Adjusted for HEI and dietary factors^e^ |  | 1.00 (ref) | 0.97 (0.91 to 1.03) | 0.96 (0.90 to 1.02) | 0.93 (0.87 to 0.99) | 0.91 (0.85 to 0.97) | .002 |
| Adjusted for BMI^f^ |  | 1.00 (ref) | 0.97 (0.91 to 1.03) | 0.97 (0.92 to 1.04) | 0.97 (0.91 to 1.03) | 1.00 (0.94 to 1.06) | .92 |
| Distal colon |  |  |  |  |  |  |  |
| No. of cases | 2633 | 527 | 481 | 529 | 540 | 556 |  |
| Base model hazard ratio (95% CI)^c^ |  | 1.00 (ref) | 0.91 (0.81 to 1.03) | 1.00 (0.88 to 1.13) | 1.02 (0.90 to 1.15) | 1.06 (0.94 to 1.19) | .13 |
| Multivariable-adjusted hazard ratio^d^ (95% CI) |  | 1.00 (ref) | 0.91 (0.81 to 1.03) | 0.99 (0.88 to 1.12) | 1.00 (0.88 to 1.13) | 0.99 (0.88 to 1.12) | .66 |
| Adjusted for HEI and dietary factors^e^ |  | 1.00 (ref) | 0.90 (0.79 to 1.02) | 0.95 (0.84 to 1.08) | 0.93 (0.82 to 1.06) | 0.88 (0.77 to 1.00) | .11 |
| Adjusted for BMI^f^ |  | 1.00 (ref) | 0.91 (0.80 to 1.03) | 0.98 (0.87 to 1.11) | 1.00 (0.88 to 1.12) | 1.00 (0.88 to 1.13) | .60 |
| Proximal colon |  |  |  |  |  |  |  |
| No. of cases | 4787 | 914 | 979 | 940 | 936 | 1018 |  |
| Base model hazard ratio (95% CI)^c^ |  | 1.00 (ref) | 1.08 (0.99 to 1.18) | 1.03 (0.94 to 1.13) | 1.03 (0.94 to 1.13) | 1.14 (1.04 to 1.25) | .02 |
| Multivariable-adjusted hazard ratio^d^ (95% CI) |  | 1.00 (ref) | 1.06 (0.96 to 1.16) | 1.00 (0.91 to 1.09) | 0.99 (0.90 to 1.08) | 1.07 (0.98 to 1.17) | .40 |
| Adjusted for HEI and dietary factors^e^ |  | 1.00 (ref) | 1.05 (0.96 to 1.15) | 0.98 (0.90 to 1.08) | 0.96 (0.87 to 1.06) | 1.00 (0.91 to 1.10) | .50 |
| Adjusted for BMI^f^ |  | 1.00 (ref) | 1.05 (0.96 to 1.15) | 0.99 (0.91 to 1.09) | 0.98 (0.90 to 1.08) | 1.07 (0.98 to 1.18) | .37 |
| Rectal |  |  |  |  |  |  |  |
| No. of cases | 2389 | 505 | 460 | 483 | 479 | 462 |  |
| Base model hazard ratio (95% CI)^c^ |  | 1.00 (ref) | 0.91 (0.80 to 1.03) | 0.95 (0.84 to 1.08) | 0.94 (0.83 to 1.07) | 0.92 (0.81 to 1.04) | .31 |
| Multivariable-adjusted hazard ratio^d^ (95% CI) |  | 1.00 (ref) | 0.90 (0.79 to 1.02) | 0.93 (0.82 to 1.06) | 0.92 (0.81 to 1.04) | 0.86 (0.76 to 0.98) | .05 |
| Adjusted for HEI and dietary factors^e^ |  | 1.00 (ref) | 0.89 (0.79 to 1.02) | 0.91 (0.80 to 1.04) | 0.87 (0.76 to 0.99) | 0.76 (0.67 to 0.88) | <.001 |
| Adjusted for BMI^f^ |  | 1.00 (ref) | 0.90 (0.79 to 1.02) | 0.93 (0.82 to 1.06) | 0.92 (0.81 to 1.04) | 0.86 (0.76 to 0.98) | .05 |

Abbreviations: HR denotes hazard ratio; CI denotes confidence interval; BMI is body mass index, calculated as weight in kilograms divided by height in meters squared; HEI is Healthy Eating Index 2015.

^a^Cases with cancer sites of C188, C189, and C260 are included in the overall analysis of 10075 cases but are censored at diagnosis date in the anatomic location analysis.

^b^Each quintile was assigned to its median value and treated as a continuous variable.

^c^Adjusted for age in years (underlying time metric), sex (male/female), and total daily energy (kcal/day) for nutrient density adjustment.

^d^Multivariable estimates were adjusted for age in years (underlying time metric), total daily energy (kcal/day), sex (male/female), race/ethnicity (American Indian/Alaskan Native, Asian, Hispanic, Non-Hispanic Black, Non-Hispanic White, Pacific Islander, Unknown), smoking by intensity (cigarettes per day: 1-10, 11-20, 21-30, 31-40, 41-60, 61+) and time since cessation (10+ years ago, 5-9 years ago, 1-4 years ago, within the last year), education level (11 years or less; 12 years, completed high school, or GED; post-high school training; some college; college and post graduate; unknown), physical activity level (never/rarely, low, moderate, high, unknown), alcohol intake (0 drinks/day, < 1 drink/day, 1-2 drinks/day, 3-4 drinks/day, 5 or more drinks/day), family history of cancer (yes/no), and self-reported health status (excellent, very good, good, fair).

^e^Multivariable model further adjusted for Healthy Eating Index (HEI)-2015 score quartiles (males - quartile 1: 21.5 to <60.6, quartile 2: 60.6 to <67.7, quartile 3: 67.7 to <74.0, quartile 4: 74.0 to 97.5; females - quartile 1: 25.9 to <63.0, quartile 2: 63.0 to <69.8, quartile 3: 69.8 to <75.6, quartile 4: 75.6 to 95.8), dietary calcium quintiles (males - quintile 1: 79.2 to <303.6, quintile 2: 303.6 to <367.0, quintile 3: 367.0 to <438.5, quintile 4: 438.5 to <551.3, quintile 5: 551.3 to 2312.8 mg/1000 kcal/day; females - quintile 1: 53.9 to <318.1, quintile 2: 318.1 to <392.6, quintile 3: 392.6 to <476.6, quintile 4: 476.6 to <605.2, quintile 5: 605.2 to 2321.7 mg/1000 kcal/day), and dietary fiber quintiles (males - quintile 1: 0.56 to <7.93, quintile 2: 7.93 to <9.69, quintile 3: 9.69 to <11.44, quintile 4: 11.44 to <13.89, quintile 5: 13.89 to 51.98 g/1000 kcal/day; females - quintile 1: 1.01 to <8.57, quintile 2: 8.57 to <10.47, quintile 3: 10.47 to <12.34, quintile 4: 12.34 to <14.93, quintile 5: 14.93 to 57.17 g/1000 kcal/day).

^f^Multivariable model further adjusted for body mass index category (<18.5 kg/m2, 18.5 to <25 kg/m2, 25 to <30 kg/m2, ≥30 kg/m2, unknown).

Supplementary Table 8. Association of UPF intake (sex-specific quintiles of % grams/day) with colorectal cancer risk in the NIH-AARP Diet and Health Study (N=461,682)

| Cancer type | N^a^ | Q1 HR (95% CI)  (N=92337) | Q2 HR (95% CI)  (N=92336) | Q3 HR (95% CI)  (N=92336) | Q4 HR (95% CI)  (N=92336) | Q5 HR (95% CI)  (N=92337) | *P* for trend^b^ |
| --- | --- | --- | --- | --- | --- | --- | --- |
| Colorectal cancer |  |  |  |  |  |  |  |
| No. of cases | 10075 | 2084 | 1947 | 1963 | 2030 | 2051 |  |
| Base model hazard ratio (95% CI)^c^ |  | 1.00 (ref) | 0.93 (0.87 to 0.99) | 0.94 (0.88 to 1.00) | 0.98 (0.92 to 1.05) | 1.03 (0.97 to 1.10) | .02 |
| Multivariable-adjusted hazard ratio^d^ (95% CI) |  | 1.00 (ref) | 0.94 (0.88 to 1.00) | 0.95 (0.89 to 1.01) | 0.98 (0.92 to 1.05) | 1.01 (0.95 to 1.07) | .21 |
| Adjusted for HEI and dietary factors^e^ |  | 1.00 (ref) | 0.93 (0.87 to 0.99) | 0.93 (0.87 to 0.99) | 0.95 (0.89 to 1.01) | 0.94 (0.88 to 1.01) | .34 |
| Adjusted for BMI^f^ |  | 1.00 (ref) | 0.94 (0.88 to 1.00) | 0.94 (0.89 to 1.01) | 0.97 (0.91 to 1.04) | 0.99 (0.93 to 1.05) | .56 |
| Distal colon |  |  |  |  |  |  |  |
| No. of cases | 2633 | 546 | 492 | 514 | 549 | 532 |  |
| Base model hazard ratio (95% CI)^c^ |  | 1.00 (ref) | 0.89 (0.78 to 1.00) | 0.92 (0.82 to 1.04) | 0.99 (0.88 to 1.12) | 0.98 (0.87 to 1.11) | .51 |
| Multivariable-adjusted hazard ratio^d^ (95% CI) |  | 1.00 (ref) | 0.90 (0.80 to 1.02) | 0.93 (0.83 to 1.06) | 0.98 (0.87 to 1.11) | 0.94 (0.83 to 1.06) | .80 |
| Adjusted for HEI and dietary factors^e^ |  | 1.00 (ref) | 0.89 (0.78 to 1.00) | 0.90 (0.80 to 1.02) | 0.93 (0.82 to 1.05) | 0.85 (0.75 to 0.97) | .07 |
| Adjusted for BMI^f^ |  | 1.00 (ref) | 0.90 (0.79 to 1.01) | 0.92 (0.82 to 1.04) | 0.96 (0.85 to 1.09) | 0.91 (0.80 to 1.03) | .42 |
| Proximal colon |  |  |  |  |  |  |  |
| No. of cases | 4787 | 982 | 927 | 923 | 958 | 997 |  |
| Base model hazard ratio (95% CI)^c^ |  | 1.00 (ref) | 0.94 (0.86 to 1.03) | 0.96 (0.87 to 1.05) | 1.01 (0.93 to 1.11) | 1.12 (1.02 to 1.22) | <.001 |
| Multivariable-adjusted hazard ratio^d^ (95% CI) |  | 1.00 (ref) | 0.95 (0.86 to 1.03) | 0.95 (0.87 to 1.04) | 1.00 (0.91 to 1.09) | 1.08 (0.98 to 1.18) | .02 |
| Adjusted for HEI and dietary factors^e^ |  | 1.00 (ref) | 0.94 (0.86 to 1.03) | 0.94 (0.86 to 1.03) | 0.97 (0.89 to 1.07) | 1.03 (0.93 to 1.13) | .22 |
| Adjusted for BMI^f^ |  | 1.00 (ref) | 0.94 (0.86 to 1.03) | 0.95 (0.86 to 1.04) | 0.99 (0.90 to 1.08) | 1.06 (0.96 to 1.16) | .05 |
| Rectal |  |  |  |  |  |  |  |
| No. of cases | 2389 | 492 | 472 | 484 | 474 | 467 |  |
| Base model hazard ratio (95% CI)^c^ |  | 1.00 (ref) | 0.95 (0.83 to 1.07) | 0.97 (0.85 to 1.10) | 0.95 (0.84 to 1.08) | 0.96 (0.84 to 1.09) | .65 |
| Multivariable-adjusted hazard ratio^d^ (95% CI) |  | 1.00 (ref) | 0.97 (0.86 to 1.11) | 1.00 (0.88 to 1.14) | 0.99 (0.87 to 1.12) | 0.98 (0.86 to 1.11) | .77 |
| Adjusted for HEI and dietary factors^e^ |  | 1.00 (ref) | 0.96 (0.85 to 1.10) | 0.98 (0.86 to 1.11) | 0.94 (0.83 to 1.07) | 0.90 (0.79 to 1.03) | .11 |
| Adjusted for BMI^f^ |  | 1.00 (ref) | 0.98 (0.86 to 1.11) | 1.00 (0.88 to 1.14) | 0.98 (0.86 to 1.12) | 0.97 (0.85 to 1.11) | .70 |

Abbreviations: HR denotes hazard ratio; CI denotes confidence interval; BMI is body mass index, calculated as weight in kilograms divided by height in meters squared; HEI is Healthy Eating Index 2015.

^a^Cases with cancer sites of C188, C189, and C260 are included in the overall analysis of 10075 cases but are censored at diagnosis date in the anatomic location analysis.

^b^Each quintile was assigned to its median value and treated as a continuous variable.

^c^Adjusted for age in years (underlying time metric), sex (male/female), and total daily energy (kcal/day) for nutrient density adjustment.

^d^Multivariable estimates were adjusted for age in years (underlying time metric), total daily energy (kcal/day), sex (male/female), race/ethnicity (American Indian/Alaskan Native, Asian, Hispanic, Non-Hispanic Black, Non-Hispanic White, Pacific Islander, Unknown), smoking by intensity (cigarettes per day: 1-10, 11-20, 21-30, 31-40, 41-60, 61+) and time since cessation (10+ years ago, 5-9 years ago, 1-4 years ago, within the last year), education level (11 years or less; 12 years, completed high school, or GED; post-high school training; some college; college and post graduate; unknown), physical activity level (never/rarely, low, moderate, high, unknown), alcohol intake (0 drinks/day, < 1 drink/day, 1-2 drinks/day, 3-4 drinks/day, 5 or more drinks/day), family history of cancer (yes/no), and self-reported health status (excellent, very good, good, fair).

^e^Multivariable model further adjusted for Healthy Eating Index (HEI)-2015 score quartiles (males - quartile 1: 21.5 to <60.6, quartile 2: 60.6 to <67.7, quartile 3: 67.7 to <74.0, quartile 4: 74.0 to 97.5; females - quartile 1: 25.9 to <63.0, quartile 2: 63.0 to <69.8, quartile 3: 69.8 to <75.6, quartile 4: 75.6 to 95.8), dietary calcium quintiles (males - quintile 1: 79.2 to <303.6, quintile 2: 303.6 to <367.0, quintile 3: 367.0 to <438.5, quintile 4: 438.5 to <551.3, quintile 5: 551.3 to 2312.8 mg/1000 kcal/day; females - quintile 1: 53.9 to <318.1, quintile 2: 318.1 to <392.6, quintile 3: 392.6 to <476.6, quintile 4: 476.6 to <605.2, quintile 5: 605.2 to 2321.7 mg/1000 kcal/day), and dietary fiber quintiles (males - quintile 1: 0.56 to <7.93, quintile 2: 7.93 to <9.69, quintile 3: 9.69 to <11.44, quintile 4: 11.44 to <13.89, quintile 5: 13.89 to 51.98 g/1000 kcal/day; females - quintile 1: 1.01 to <8.57, quintile 2: 8.57 to <10.47, quintile 3: 10.47 to <12.34, quintile 4: 12.34 to <14.93, quintile 5: 14.93 to 57.17 g/1000 kcal/day).

^f^Multivariable model further adjusted for body mass index category (<18.5 kg/m2, 18.5 to <25 kg/m2, 25 to <30 kg/m2, ≥30 kg/m2, unknown).

Supplementary figure 1. Association of ultra-processed food intake (quintiles of nutrient-adjusted g/1000 kcal/day) with colorectal cancer risk overall and by subgroup

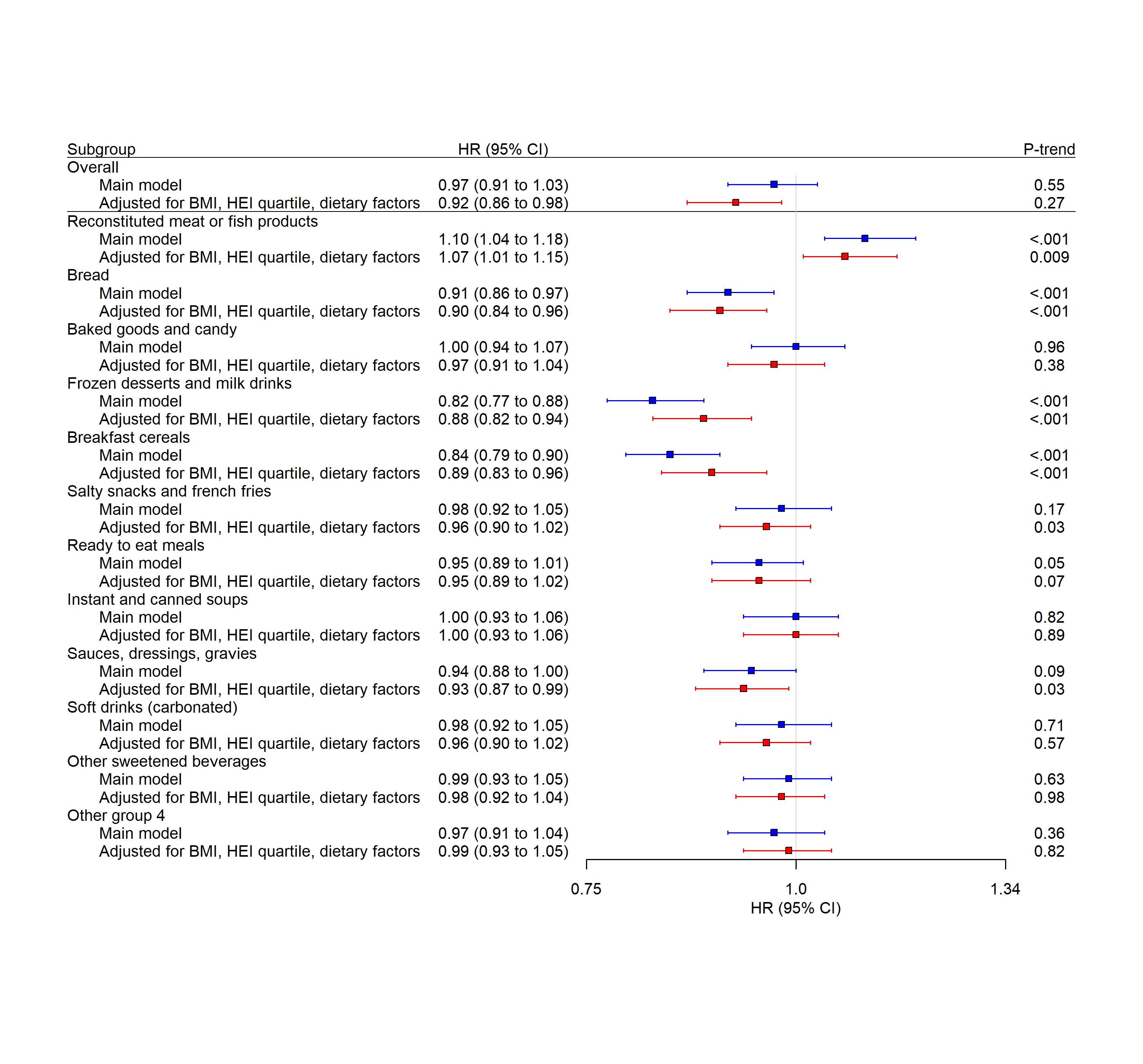

Abbreviations: UPF – Ultra-processed food; BMI – body mass index; HEI – Healthy Eating Index 2015; HR – hazard ratio; CI – confidence interval

Hazard ratios are for quintile 5 of grams of UPF subgroup with reference to quintile 1. HRs were estimated using a Cox proportional hazard regression model with age as the underlying time metric adjusted total daily energy (kcal/day), sex (male/female), race/ethnicity (American Indian/Alaskan Native, Asian, Hispanic, Non-Hispanic Black, Non-Hispanic White, Pacific Islander, Unknown), smoking by intensity (cigarettes per day: 1-10, 11-20, 21-30, 31-40, 41-60, >60) and time since cessation (≥10 years ago, 5-9 years ago, 1-4 years ago, within the last year), education level (11 years or less; 12 years, completed high school, or GED; post-high school training; some college; college and post graduate; unknown), physical activity level (never/rarely, low, moderate, high, unknown), alcohol intake (0 drinks/day, < 1 drink/day, 1-2 drinks/day, 3-4 drinks/day, ≥ 5 drinks/day), family history of cancer (yes/no), self-reported health status (excellent, very good, good, fair, unknown), and all other UPF subgroups. Dietary factors included nutrient-density adjusted quintiles of dietary calcium and nutrient-density adjusted quintiles of dietary fiber. P-trend was estimated by assigning each quintile to its median value and treating it as a continuous variable.
